# The feasibility and efficacy of a virtual, symptom-guided aerobic exercise intervention to improve cognition in mild traumatic brain injury: A single-blind pilot randomized control trial with an active comparator group

**DOI:** 10.64898/2025.12.11.25342088

**Authors:** Emma M. Tinney, Mark C. Nwakamma, Goretti España-Irla, Lauren Kong, Colette Chen, Jeremy Hwang, Amanda O’Brien, Madeleine Perko, Ryan Luke Sodemann, Jacqueline Caefer, Julia Manczurowsky, Alexandra Stillman, Charles H. Hillman, Timothy P. Morris

**Author notes:** Corresponding Author: Emma M. Tinney.

## Abstract

**Background:** Mild traumatic brain injury (mTBI) affects millions worldwide, with cognitive impairment substantially impacting daily functioning. Despite this burden, evidence-based non-pharmaceutical interventions remain lacking in clinical practice. Emerging evidence suggests aerobic exercise may improve post-mTBI cognition; however, the methodological limitations, particularly inadequate control groups, prevent definitive conclusions.

**Objective:** This pilot randomized controlled trial examined the feasibility and preliminary efficacy of a 12-week virtual exercise intervention for community-dwelling adults aged 18-55 years with mTBI within one year of injury.

**Methods:** Thirty-seven participants were randomized to either symptom-guided aerobic exercise or active balance control; both delivered virtually three times weekly for 30 minutes over 12 weeks. Primary outcomes assessed feasibility metrics; secondary outcomes examined cognitive function.

**Results:** Of enrolled participants, 75% completed the intervention with 94.2% session adherence and zero adverse events, demonstrating excellent feasibility and safety. The aerobic group demonstrated greater improvements in executive function compared to balance controls, with large effect sizes for TMT B-A difference scores in both post-intervention comparisons (Hedges’ *g* = 1.20, 95% CI [0.00, 2.41]) and Group × Time interactions (Hedges’ *g* = 1.38, 95% CI [0.27, 2.49]). Additionally, the aerobic group reported fewer sleep disturbances post intervention (*g* = 1.65, 95% CI [0.22, 3.09]).

**Conclusion:** These findings establish that virtual, supervised, symptom-guided exercise interventions are feasible and safe for mTBI populations, with preliminary evidence suggesting aerobic exercise specifically benefits cognitive flexibility and sleep quality following mTBI. A fully powered randomized controlled trial is warranted to confirm these effects

## Introduction

Mild traumatic brain injury (mTBI) affects an estimated 60 million individuals worldwide annually, with a substantial proportion experiencing persistent cognitive impairment that significantly impacts return to work and daily functioning^1^. Despite this considerable public health burden, there are no FDA-approved treatments to directly address cognitive dysfunction following mTBI^2^, leaving clinicians with limited options beyond symptom monitoring and activity modification.

Aerobic exercise has emerged as a promising therapeutic approach, with preclinical evidence demonstrating neuroprotective effects, including reduced neuroinflammation, enhanced neurogenesis, and improved synaptic plasticity^3^. Previous human studies examining exercise interventions for cognitive recovery after mTBI have been limited by significant heterogeneity, including inadequate control groups, unsupervised interventions, and lack of individualized symptom-guided prescriptions^4–12^. These limitations have prevented definitive conclusions about exercise efficacy and optimal implementation approaches for mTBI populations.

The current study addresses critical gaps in prior studies through three key innovations. First, we implemented a virtual yet supervised exercise intervention that overcomes barriers to exercise participation in mTBI populations, such as transportation challenges, symptom exacerbation in gym environments, and cognitive demands of navigating unfamiliar spaces and scheduling^13^. Second, we implemented a symptom-threshold approach using metrics from validated exercise tolerance testing protocols^14^, allowing for real time modification of intensity based on individual symptoms. This personalized approach minimizes risk of symptom exacerbation while promoting self-efficacy and adherence. Third, we included an active balance control group to isolate aerobic exercise effects from standard practice of care and general physical activity benefits, a methodological control absent from prior studies. Therefore, the purpose of this study was to test the feasibility of an individualized, virtual, supervised exercise intervention in mTBI.

The primary aim of this pilot study was to establish feasibility of delivering a 12-week supervised virtual exercise intervention in community-dwelling adults with a recent mTBI (within one year of injury), including recruitment rates, adherence, retention, and safety parameters. Secondary aims examined precision estimates of the effect of the intervention versus the active comparator group (means, variances, and effect sizes with 95% confidence intervals)^15,16^ for changes in cognitive function and a battery of psychosocial questionnaires.

## Methods

Details of all study protocols are described in the protocol paper^17^ and below. This study received ethical approval from the Northeastern University Institutional Review Board (#24-10-30).

### Study design

This was a two-arm randomized single-blind control trial. After baseline testing, participants were randomized after the baseline session using a block randomization in block sizes of four and six with equal allocation to each group, with allocation concealed in REDCap. Participants were blinded to study hypotheses, but investigators were not blinded. Participants engaged in supervised exercise sessions three-times per week for twelve weeks with outcome assessments obtained before and after the intervention.

### Participants

Adults aged 18-55 years with a diagnosis of mTBI within the past year were recruited for participation in an exercise intervention (The Exercise and Concussion Health Study (TECHS)) (ClinicalTrials.gov: NCT06494592) via advertisements around the Greater Boston area. Eligibility required that all participants received a formal concussion or mTBI diagnosis by a physician (AS). Diagnoses were confirmed using OSU-TBI-ID questionnaire^18^, requiring a direct blow or impact to the head with either a loss of consciousness of <5 minutes, dazed, or a gap in memory for <5 minutes. Participants were excluded if they had a skull breach, subdural hematoma, prior diagnosis of cognitive or physical disability, clinical diagnosis of neurological or neuropsychiatric disorder, undergoing treatment for cardiovascular events, not fluent in English, not medically cleared for exercise, use of an assisted walking device, or not magnetic resonance imaging compatible.

Between January 2023 and June 2025 216 inquiries were received, 83 individuals were assessed for eligibility. 37 were consented and enrolled into the study. See the consort diagram in Figure 1 for details. An a-priori sample size of 12 per arm was chosen based on Julious et al., 2005, considering the feasibility of patient recruitment and the optimal precision and variance about the effect estimates to inform future studies.

**Figure 1:**
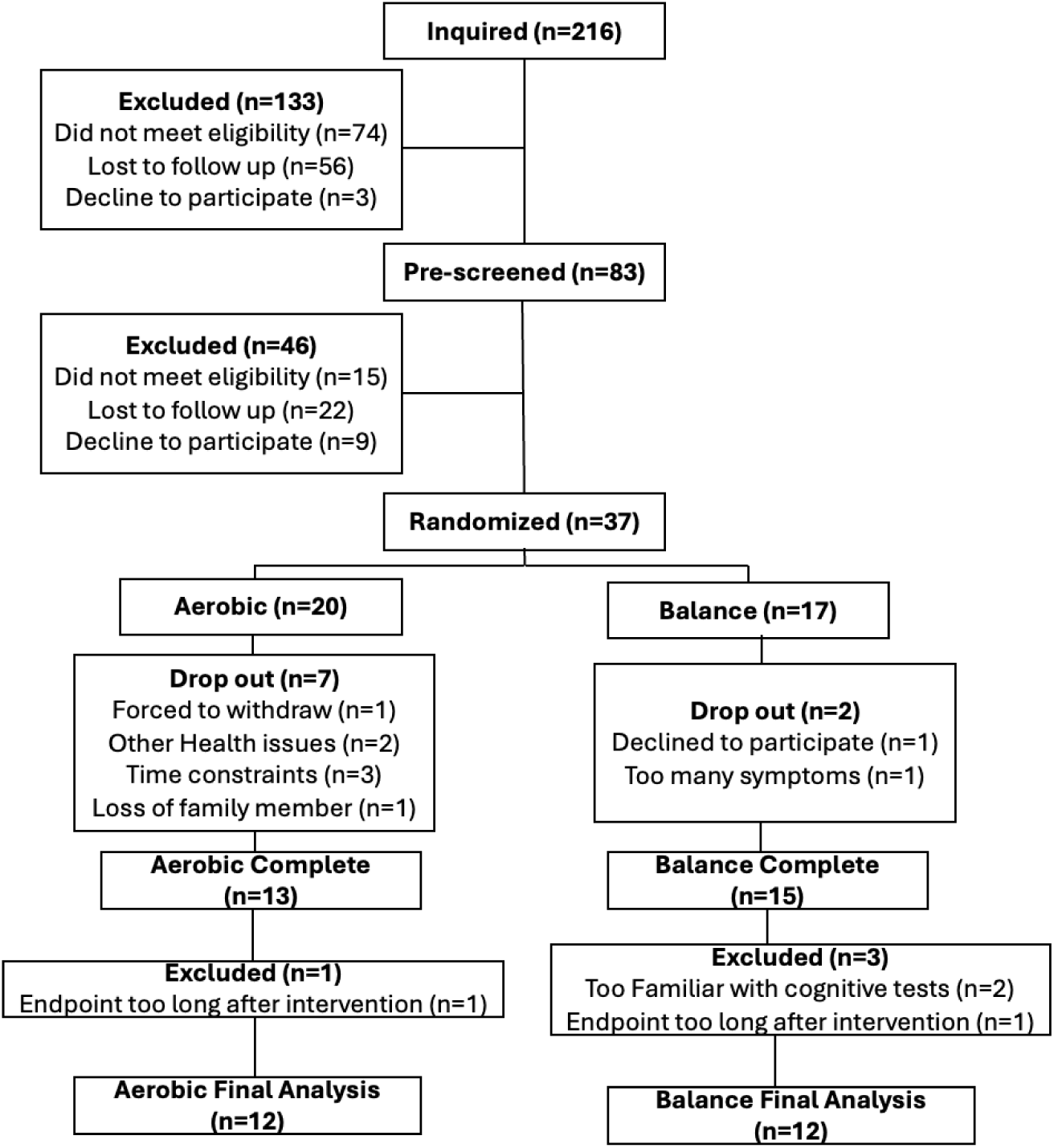
Consort diagram.

### Cognitive Testing

Trail Making Test^19^, Verbal Fluency^20^, and Hopkins Verbal Learning Test^21^ were administered at baseline and endpoint. These were chosen to provide measures of domains with known deficits after a mTBI and are commonly used in clinical practice. All cognitive testing was administered by two trained research staff (EMT and MCN).

### Symptom Threshold and Cardiorespiratory Fitness Testing

We utilized an in person hybrid modified exertional bike test, similar to the Buffalo concussion treadmill test and others^14,22^ to objectively determine exercise intensity (via heart rate) at which concussion symptoms increase ≥3 units above baseline, combined with sub-maximal VO max estimation using the Astrand-Rhyming protocol^23^ (Figure 2). The assessment consisted of three phases on a cycle ergometer: (1) 7-minute self-paced warm-up, (2) 6-minute Astrand-Rhyming test at 50 RPM with predetermined workload to estimate VO max, and (3) symptom threshold test with resistance increasing every 30 seconds until meeting termination criteria (≥3-point symptom increase, age-predicted maximum heart rate, or RPE ≥18). Heart rate (Polar H7), RPE (Borg scale^24^), and symptoms were recorded every two minutes. The symptom threshold heart rate was used to prescribe aerobic exercise intensity for intervention sessions. Two trained staff (EMT, MCN) administered all testing at baseline and post-intervention (see Nwakamma et al., for details).

**Figure 2:**
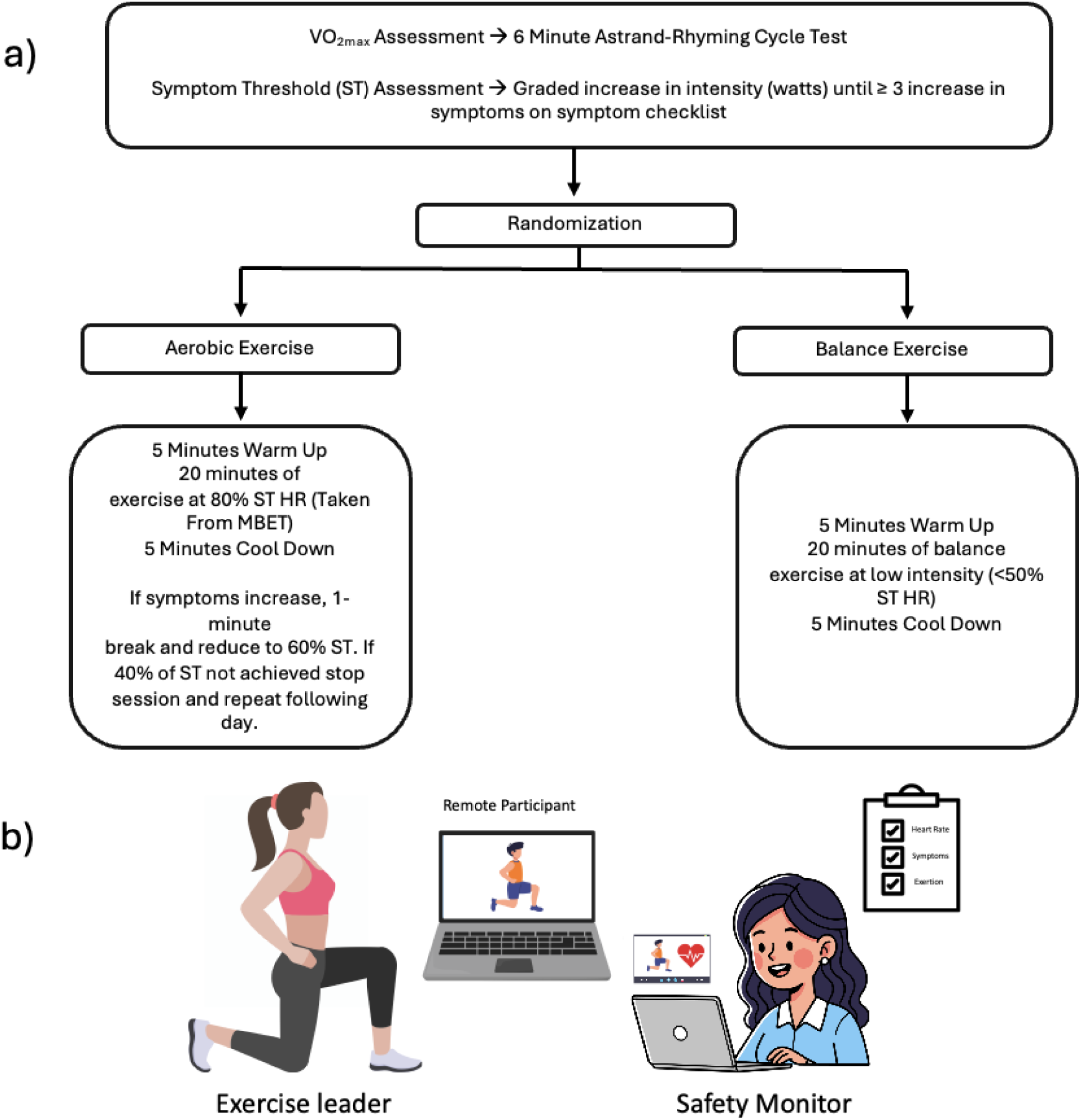
Modified Exertional Bike Test and Intervention. a) Modified Exertional Bike Test and decision tree. b) virtual intervention set up. Study staff led exercise sessions from a dedicated room in the research institute. The safety officer monitored the session for several criteria including flushed face, increased perfusion, pale features and followed an emergency protocol if signs were observed.

### Balance Test

The Balance Error Scoring System (BESS)^25^ was used to evaluate postural stability, consisting of three stance positions performed on two different surfaces (firm and foam), yielding six total testing conditions held for 20 seconds. The BESS was administered and scored by two trained research staff (EMT and MCN).

### Psychosocial Assessments

A battery of questionnaires was administered at baseline and endpoint including the Pittsburgh Sleep Quality Index (PSQI)^26^, Patient-Reported Outcomes Measurement Information System (PROMIS)^27–29^, and International Physical Activity Questionnaire (IPAQ)^30^, asking specifically about pre-injury physical activity engagement.

### Intervention

All participants completed supervised one-on-one virtual exercise sessions three times weekly for 30 minutes over 12 weeks (36 total sessions). Weekly texts confirmed scheduling, adherence, and remaining sessions. Each intervention group had three sets of five exercises with modifications to maintain engagement and progression. Two trained staff led each virtual session: one exercise leader and one safety observer. Heart rate (HR) was monitored via hyperate4health (https://hyperate4health.netlify.app/) and manually recorded every 5 minutes (continuous recording worked for only 54% of participants for ≥18 sessions). Participants rated symptoms and exertion every five minutes. Aerobic sessions targeted 20 minutes at 65-80% of symptom threshold HR. If symptoms increased by ≥3 points, participants took a one-minute break and reduced intensity to 60% of threshold HR. If symptoms persisted, another break occurred and intensity decreased to 40% of threshold HR (Figure 2).

### Endpoint satisfaction survey

Participants were asked to complete an optional satisfaction survey, answering Likert scale questions about scheduling, communication, difficulty, and open-ended responses about satisfaction with the study.

### Statistical Analysis

Descriptive statistics are provided for all feasibility metrics. To calculate demographic differences between groups, we performed analyses of variance (ANOVA), reporting sum of squares, and 95% confidence intervals. For categorical variables, we performed chi-squared tests, reporting χ² and 95% confidence intervals.

Statistical models to test for differences in outcomes at end point and rate of change over time between groups were used. However, inferences were not made based on p-values as sample size was chosen to demonstrate feasibility, precision, and variance around the mean difference. Therefore, all model results are reported as effect sizes and 95% confidence intervals around the point estimate.

We used two complementary approaches to evaluate treatment effects. All models controlled for four fixed covariates: age, sex, education, and days since injury. Forward selection with AIC identified additional covariates improving model fit (previous injuries, adherence rate, baseline VO max, baseline symptoms, total METs, time in heart rate zone). VO max analyses excluded baseline VO max, and symptom analyses excluded baseline symptoms to avoid circularity.

We conducted Analysis of Covariance (ANCOVA) to compare post-intervention scores between groups while controlling for baseline performance. Assumptions were verified using Shapiro-Wilk, Levene’s, and Group × Pre-Score interaction tests. Outliers were identified but not removed. We also fitted linear mixed-effects models to assess group differences in change over time. Models used restricted maximum likelihood estimation (*lme4* package in R). Full details are in supplementary material 1.

## Results

### Participants

Thirty-seven participants signed informed consent, underwent baseline testing and were randomized to one of the two study arms. Nine participants withdrew from the study before completing the intervention (24%) and four participants were excluded from the final analysis for varying reasons (Figure 1). The final sample included 24 participants, 12 in each group. Groups did not differ in demographics except for biological sex (Table 1).

**Table 1.**
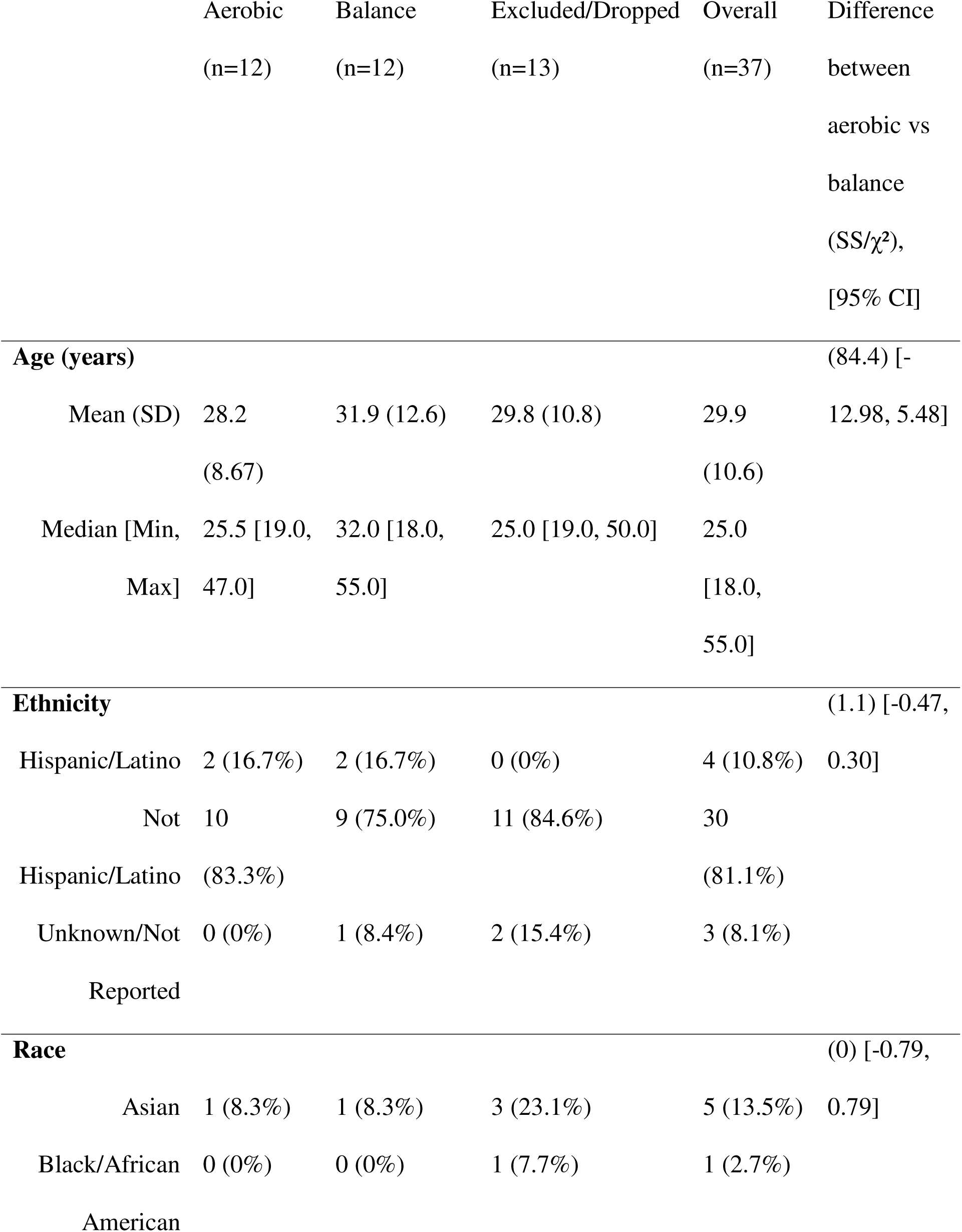
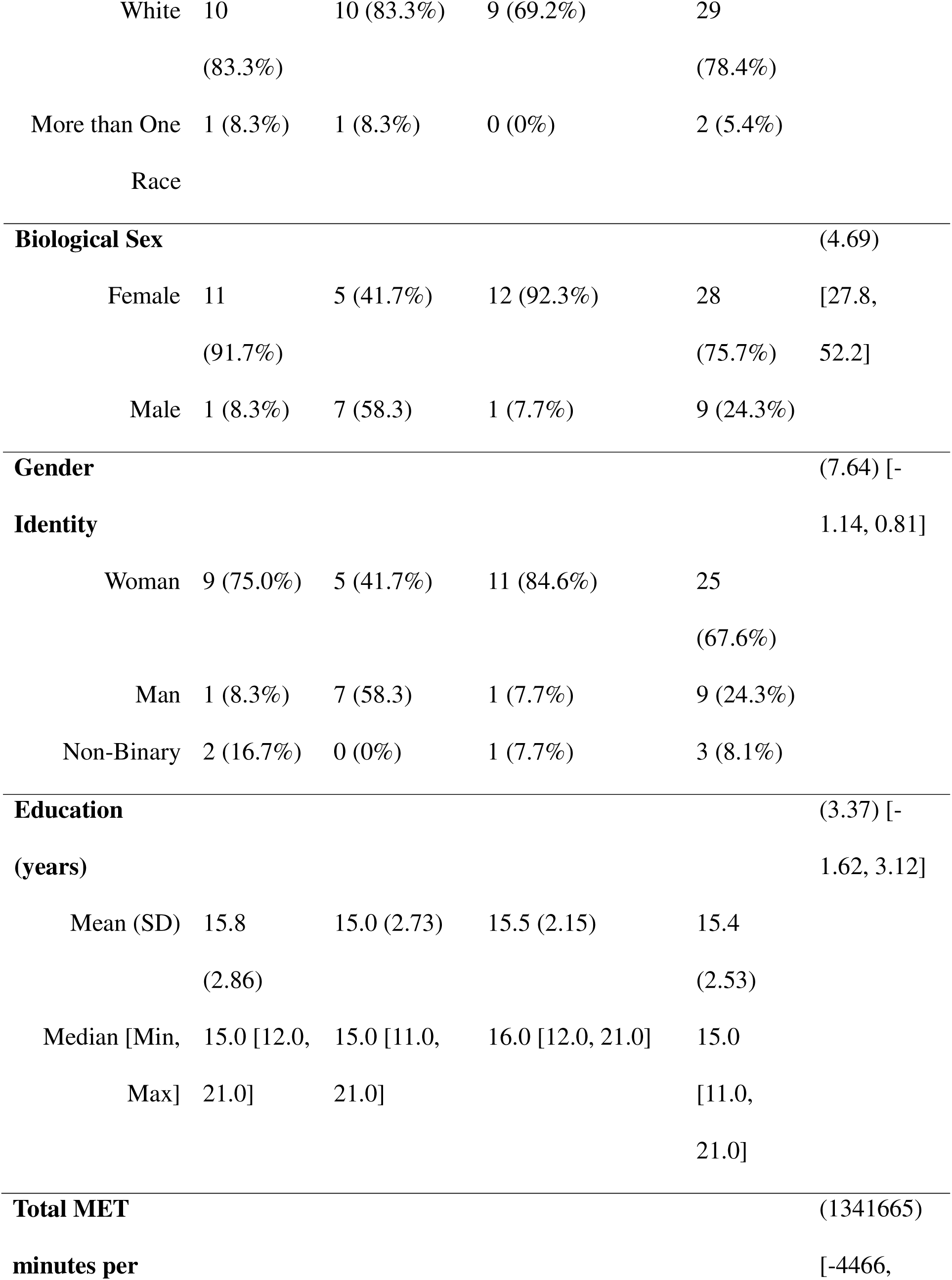
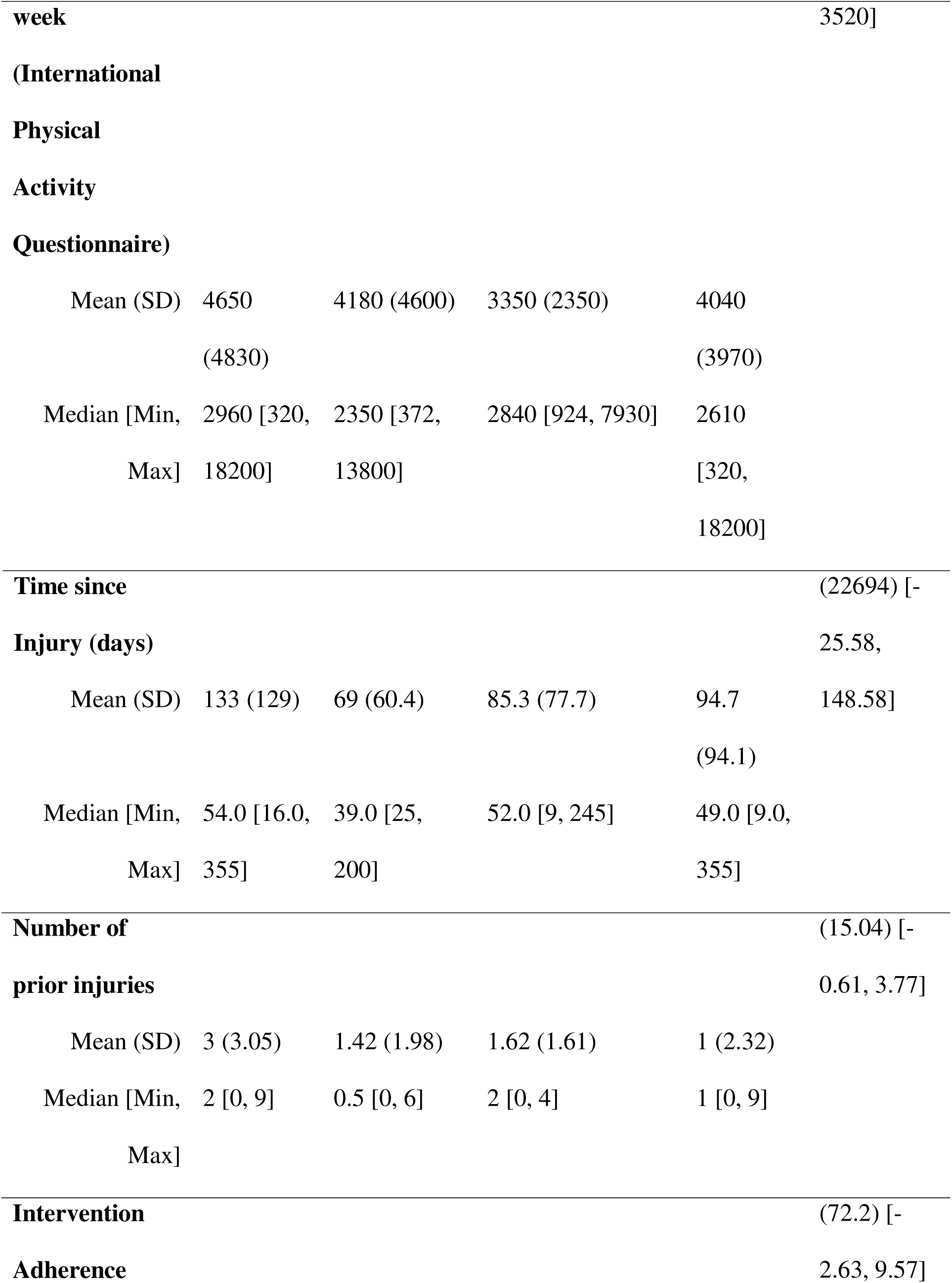
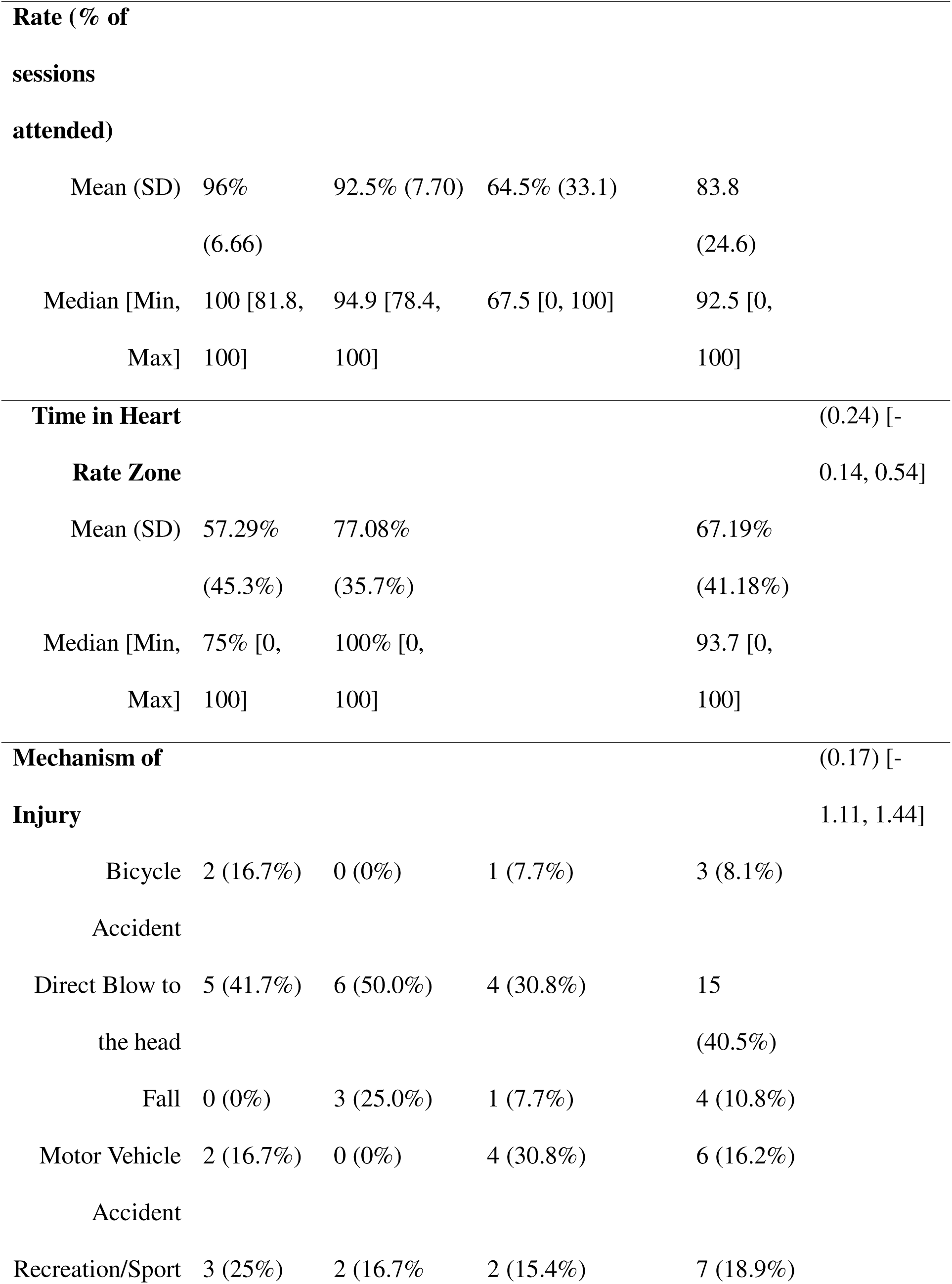
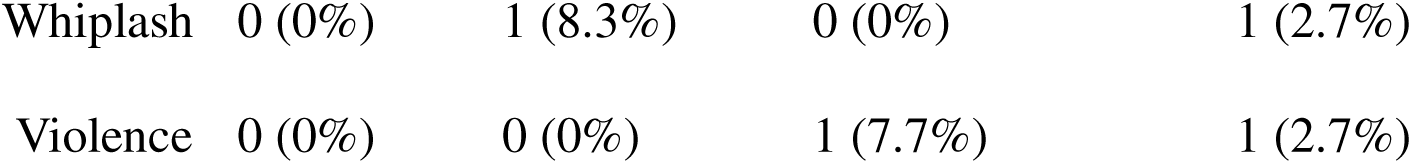
Participant Demographics.

### Adverse Events

No participant experienced any adverse events during the baseline, endpoint, or intervention. However, several minor events happened outside the study that affected participation in the study. One participant was forced to withdraw due to diagnosis of postural orthostatic tachycardia syndrome, but this was not due to the intervention. One participant developed pneumonia but received a doctor’s clearance to continue participating and resumed full protocol. One participant dropped out due to receiving unspecified inpatient care.

### Intervention feasibility

In the aerobic exercise intervention (n=12), participants were exercised at 65-80% of their symptom-threshold heart rate for 36 sessions with no adverse events. Seven of the twelve participants exceeded their prescribed HR zones for at least 50% of the intervention, but none experienced clinically significant symptom exacerbation (mean three-unit increase). Notably, 10 of 12 participants demonstrated decreased correlations between heart rate and symptom severity over time, indicating improved exercise tolerance (supplementary material 10). The balance intervention successfully minimized increases in HR with clear numerical differences in HR between intervention arms (Supplementary Material 14). An average adherence rate of 96% was achieved for the aerobic condition and 92.5% for balance. Participants’ reported a three-unit exercise-induced symptom severity increase in 3.8% of all completed sessions. Twenty-one out of 24 (87.5%) participants reported at least one symptom on the CIF. (Supplementary Material 3).

### Endpoint Differences of Primary outcomes

Pre-post changes in primary cognitive and behavioral outcomes are reported as mean and standard deviation (Table 2).The aerobic group reported fewer sleep disturbances compared to the balance group after controlling for baseline sleep quality and total physical activity levels (β=3.75,SE=1.46,*g*=1.65,95%CI[0.22, 3.09]) (Figure 3c). Specifically, the aerobic group displayed a 3.75-unit greater reduction in sleep disturbance scores. The aerobic group completed the TMT-B an average of 13.15 seconds faster than the balance group after controlling for baseline performance and number of prior injuries (β=13.15,SE= 6.05,g=1.24,95%CI[0.04,2.45]) (Figure 3c). The aerobic group demonstrated greater improvement in the TMT B-A difference score compared to the balance group after controlling for baseline performance and number of prior injuries (β=13.36,SE=6.31,*g*=1.20,95% CI[0.00,2.41]). Specifically, the aerobic group showed a 13.36-second greater reduction in the time difference between TMT-B and TMT-A (Figure 3c). The balance group demonstrated higher PROMIS global physical health compared to the aerobic group after controlling for baseline performance, number of prior injuries, and time in heart rate zone (β = -7.97, SE = 3.67, g = -1.33, 95% CI [-2.63, -0.03]) (Figure 3c). Specifically, the aerobic group scored 7.97 points lower on PROMIS GPH at post-intervention compared to the balance group, representing a large effect size (g = -1.33).

**Figure 3.**
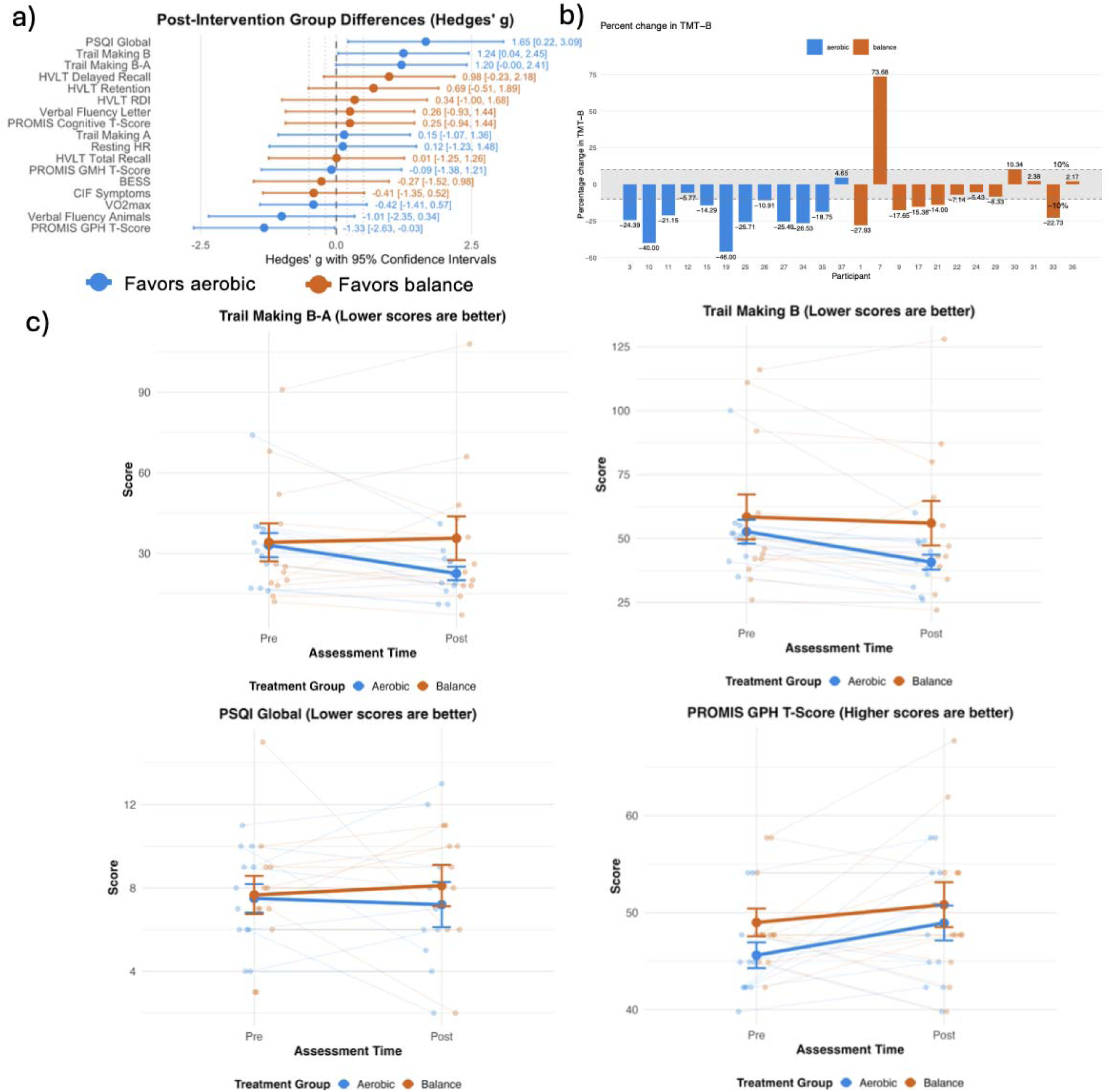
Forrest plots and Interaction plots A. a) Forest Plot of Post Intervention Group Differences b) Percent change in TMT B test, with 10% change showing clinically significant improvement. Negative percentages show faster TMT B times at endpoint. c) Pittsburg Sleep Quality Index (PSQI), Trail making test (TMT) B, Trail making test B-A score, and PROMIS Global Physical Health T Score.

**Table 2.**
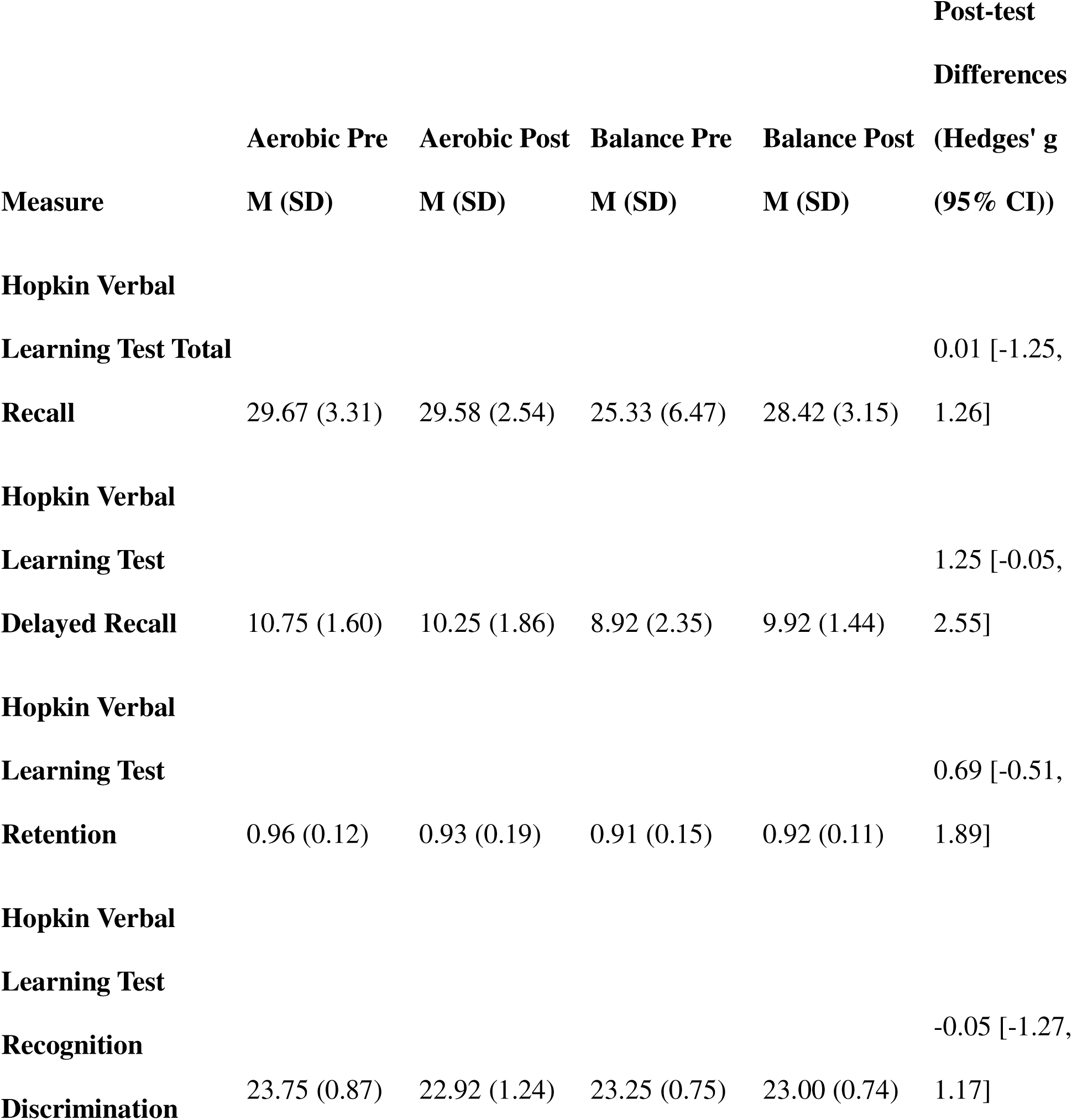
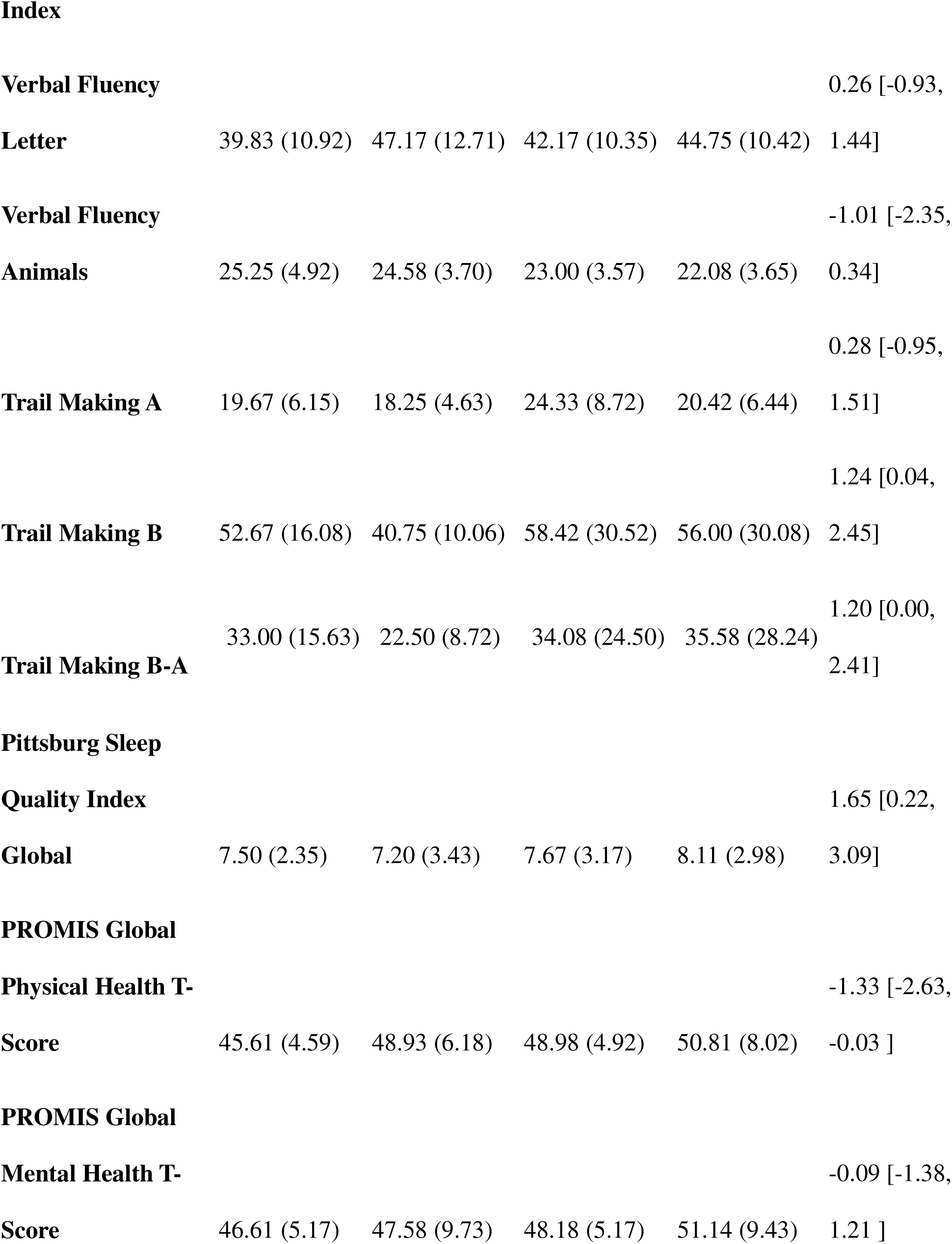
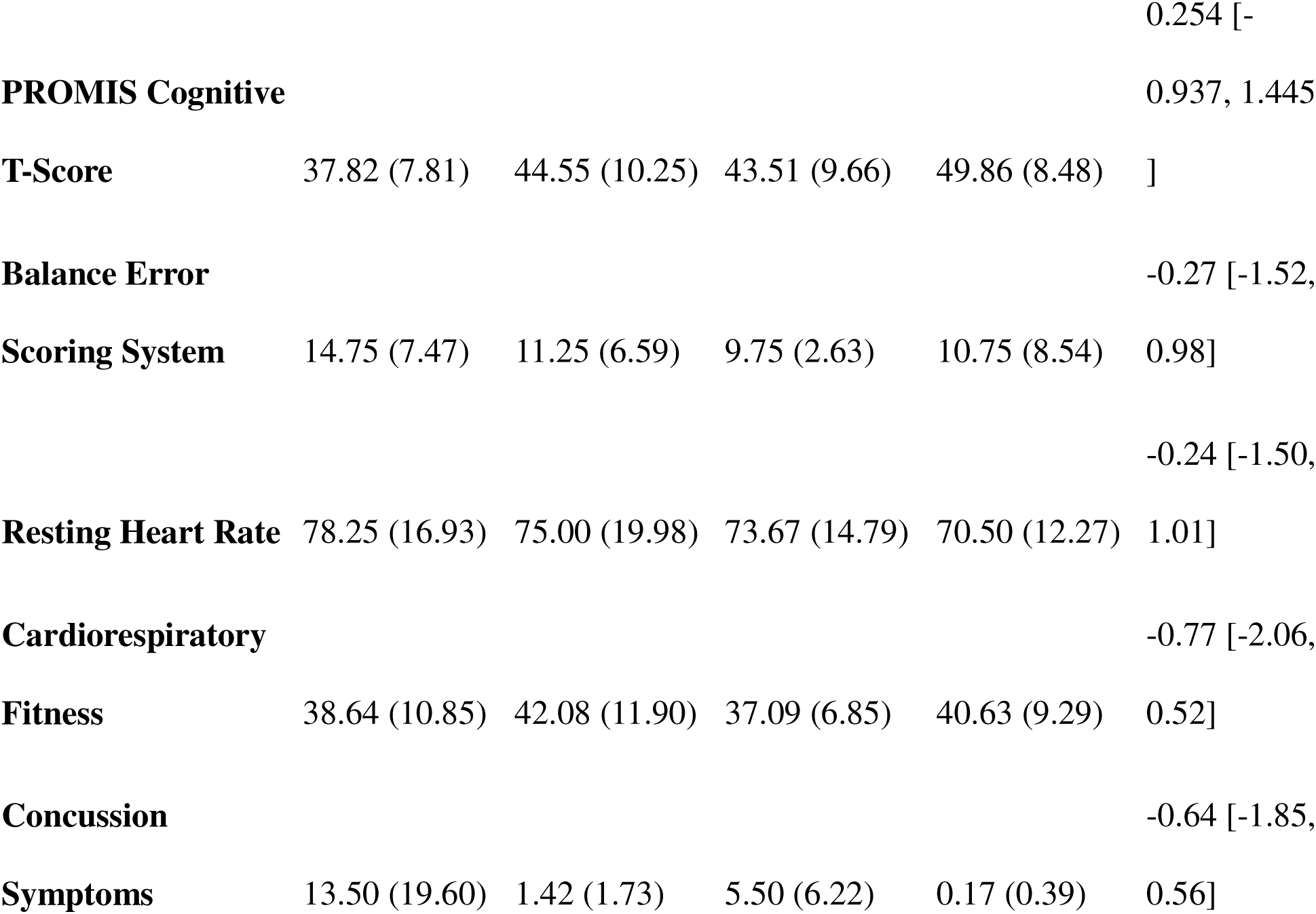
Group Means at Baseline and Endpoint.

|  |  |  |  |  | **Post-test**<br>**Differences** |
| --- | --- | --- | --- | --- | --- |
|  | **Aerobic Pre** | **Aerobic Post** | **Balance Pre** | **Balance Post** | **(Hedges' g**<br>**(95% CI))** |
| **Measure** | **M (SD)** | **M (SD)** | **M (SD)** | **M (SD)** |  |
| **Hopkin Verbal**<br>**Learning Test Total** |  |  |  |  | 0.01 [-1.25, |
| **Recall** | 29.67 (3.31) | 29.58 (2.54) | 25.33 (6.47) | 28.42 (3.15) | 1.26] |
| **Hopkin Verbal**<br>**Learning Test** |  |  |  |  | 1.25 [-0.05, |
| **Delayed Recall** | 10.75 (1.60) | 10.25 (1.86) | 8.92 (2.35) | 9.92 (1.44) | 2.55] |
| **Hopkin Verbal**<br>**Learning Test** |  |  |  |  | 0.69 [-0.51, |
| **Retention** | 0.96 (0.12) | 0.93 (0.19) | 0.91 (0.15) | 0.92 (0.11) | 1.89] |
| **Hopkin Verbal**<br>**Learning Test** |  |  |  |  | -0.05 [-1.27, |
| **Recognition** |  |  |  |  | 1.17] |
| **Discrimination** | 23.75 (0.87) | 22.92 (1.24) | 23.25 (0.75) | 23.00 (0.74) |  |
| Index |  |  |  |  |  |
| **Verbal Fluency** |  |  |  |  | 0.26 [-0.93, |
| **Letter** | 39.83 (10.92) | 47.17 (12.71) | 42.17 (10.35) | 44.75 (10.42) | 1.44] |
| **Verbal Fluency** |  |  |  |  | -1.01 [-2.35, |
| **Animals** | 25.25 (4.92) | 24.58 (3.70) | 23.00 (3.57) | 22.08 (3.65) | 0.34] |
|  |  |  |  |  | 0.28 [-0.95, |
| **Trail Making A** | 19.67 (6.15) | 18.25 (4.63) | 24.33 (8.72) | 20.42 (6.44) | 1.51] |
|  |  |  |  |  | 1.24 [0.04, |
| **Trail Making B** | 52.67 (16.08) | 40.75 (10.06) | 58.42 (30.52) | 56.00 (30.08) | 2.45] |
|  |  |  |  |  | 1.20 [0.00, |
| **Trail Making B-A** | 33.00 (15.63) | 22.50 (8.72) | 34.08 (24.50) | 35.58 (28.24) | 2.41] |
| **Pittsburg Sleep** |  |  |  |  |  |
| **Quality Index** |  |  |  |  | 1.65 [0.22, |
| **Global** | 7.50 (2.35) | 7.20 (3.43) | 7.67 (3.17) | 8.11 (2.98) | 3.09] |
| **PROMIS Global** |  |  |  |  |  |
| **Physical Health T-** |  |  |  |  | -1.33 [-2.63, |
| **Score** | 45.61 (4.59) | 48.93 (6.18) | 48.98 (4.92) | 50.81 (8.02) | -0.03 ] |
| **PROMIS Global** |  |  |  |  |  |
| **Mental Health T-** |  |  |  |  | -0.09 [-1.38, |
| **Score** | 46.61 (5.17) | 47.58 (9.73) | 48.18 (5.17) | 51.14 (9.43) | 1.21 ] |
|  |  |  |  |  | 0.254 [- |
| **PROMIS Cognitive** |  |  |  |  | 0.937, 1.445 |
| **T-Score** | 37.82 (7.81) | 44.55 (10.25) | 43.51 (9.66) | 49.86 (8.48) | ] |
| **Balance Error** |  |  |  |  | -0.27 [-1.52, |
| **Scoring System** | 14.75 (7.47) | 11.25 (6.59) | 9.75 (2.63) | 10.75 (8.54) | 0.98] |
|  |  |  |  |  | -0.24 [-1.50, |
| **Resting Heart Rate** | 78.25 (16.93) | 75.00 (19.98) | 73.67 (14.79) | 70.50 (12.27) | 1.01] |
| **Cardiorespiratory** |  |  |  |  | -0.77 [-2.06, |
| **Fitness** | 38.64 (10.85) | 42.08 (11.90) | 37.09 (6.85) | 40.63 (9.29) | 0.52] |
| **Concussion** |  |  |  |  | -0.64 [-1.85, |
| **Symptoms** | 13.50 (19.60) | 1.42 (1.73) | 5.50 (6.22) | 0.17 (0.39) | 0.56] |

No further large effect sizes were observed in post-test differences between groups. AIC-guided covariate selection revealed that time in heart rate zone, adherence rate, and number of prior injuries consistently improved model fit across multiple cognitive and health outcomes. However, neither time in heart rate zone nor adherence rate correlated with magnitude of cognitive or sleep improvements, suggesting that exceeding prescribed intensity thresholds may not confer additional benefits and that high adherence rates in both groups contributed equally to positive outcomes. Notably, number of prior injuries emerged as a potential moderator of treatment response, particularly for the balance group’s delayed recall performance, though small subgroup sizes warrant cautious interpretation. See Supplementary Material for complete ANCOVA results (12), outcome visualizations (4), expanded exploratory covariate analyses (5-7), and complimentary results of the linear mixed effects models assessing differences in change over time between groups (13).

### Study Feedback

27 participants (72%) completed the endpoint satisfaction survey collected via email. Generally. participants were satisfied with the study but recommended improved scheduling. Many participants expressed gratitude for the study and that participating helped in their subjective recovery (Supplementary Material 9).

## Discussion

This pilot study tested the feasibility of a single-blinded, supervised, virtual exercise intervention for community-dwelling individuals within one year of mTBI. Results demonstrated that this approach is feasible and safe, with high adherence and no adverse events. Participants reported high satisfaction with study procedures, and preliminary evidence suggests aerobic exercise may improve cognitive flexibility compared to an active balance control group mimicking typical vestibular physical therapy.

These findings demonstrate that virtual aerobic exercise can safely and feasibly be utilized as a rehabilitative intervention following mTBI, effectively overcoming traditional barriers to exercise participation in mTBI populations. The supervised format with continuous symptom monitoring enabled real-time intensity adjustments while maintaining safety and engagement. Although the dropout rate was slightly higher than other exercises studies in patient populations (between 15-20%^31,32^), this was improved throughout the study with enhanced participant communication. Zero adverse events, high adherence, and symptom exacerbation in only 3.8% of sessions demonstrate adequate safety protocols. Participants frequently exceeded prescribed heart rate zones suggesting difficulty of floor-based exercises (compared to running or cycling) to manipulate HR. However, exercising above the symptom-threshold did not increase symptoms, and the heart rate-symptom correlations decreased over time with continued exercise exposure (Supplementary Material 10). These results support that the intervention is feasible, acceptable, and safe for mTBI populations, warranting progression to a fully powered randomized controlled trial.

Improvement in TMT-B and TMT B-As in the aerobic group align with several prior studies demonstrating TMT improvements in individuals with TBI of varying severities following aerobic exercise ^12,33^, though both studies lacked a control group. This holds clinical significance given that TMT-B measures executive function, a cognitive domain typically impaired in TBI^34,35^ and corresponds with functional recovery^36,37^. The mechanisms underlying exercise related improvements in executive function may be driven by improved synaptic plasticity and associated downstream cellular and molecular mechanisms in prefrontal and subcortical networks that support executive control^39,40^. Given our balance intervention consisted of non-aerobic physical activity, which itself has shown improvement in cognitive function in other populations^41^, it is unclear why TMT-B appears particularly sensitive to aerobic exercise post-TBI. Notably, here, 83% of aerobic participants achieved clinically meaningful improvement (≥10% faster completion times) on TMT-B, compared to 42% of balance (Figure 3C). The improvements in TMT-B following aerobic exercise may have meaningful implications for real-world functional outcomes, including return to work, academic performance, and independent living skills. Prior work has shown improvement in TMT-B using deep brain stimulation in TBI^42^, while our study shows that aerobic exercise may provide a non-invasive and accessible therapeutic option. Further discussion of cognitive changes is found in supplementary material 13.

The aerobic group demonstrated fewer sleep disturbances compared to the balance group at post-intervention. This finding holds clinical significance given the persistent nature of sleep dysfunction following TBI. Sleep deficits after TBI can persist for years post-injury, representing one of the most common and debilitating complaints in TBI^43^. Beyond their immediate impact on quality of life, sleep disturbances are strongly associated with poorer cognitive outcomes and compromised brain health trajectories after TBI^44^, positioning sleep as a critical therapeutic target for recovery. The mechanisms underlying exercise-related improvements in sleep may operate through both direct and indirect pathways. Exercise directly influences circadian rhythm regulation, adenosine accumulation, and thermoregulatory processes that facilitate sleep onset and maintenance^52^. Indirectly, exercise may improve sleep by reducing anxiety and depressive symptoms, enhancing mood regulation, and promoting daytime wakefulness, all factors commonly disrupted after TBI. These converging mechanisms position aerobic exercise as a particularly promising therapeutic intervention for treating post-TBI sleep disturbances. Despite this theoretical foundation, empirical evidence supporting exercise interventions for sleep in TBI populations remains limited. In one of the few studies, Damanio and colleagues^47^ demonstrated that sleep quality improved following 8 weeks of daily aerobic exercise in chronic TBI. However, the absence of a control group limits causal inference from those findings. The current study addresses this critical gap with an active control group, providing robust evidence that aerobic exercise drives improvements in sleep.

Neither group demonstrated changes to cardiorespiratory fitness (CRF), likely because participants exercised at 65-80% of symptom threshold HR, which was lower than HR max for most participants. Importantly though, CRF increases appear unnecessary for beneficial effects on executive function in mTBI, suggesting exercise influences brain health through pathways independent of cardiovascular conditioning. Indeed, prior work has shown that light intensity exercise can increase executive function in non-injured adults ^49^. Alternative mechanisms of action may include acute exerc*ise-induced increases in cerebral blood flow and BDNF, reduced neuroinflammation, enhanced neurogenesis, and improved sleep quality, all achievable without cardiovascular conditioning^50^. These findings have significant clinical implications: symptom-threshold guided exercise can yield meaningful cognitive benefits at intensities below those needed for CRF improvements. This supports individualized, moderate-intensity exercise protocols that are safer, more tolerable, and reduce barriers to mTBI rehabilitation by minimizing concerns about symptom exacerbation.

### Limitations

Our study has several limitations. The small sample size limits generalizability, but was chosen a-priori to provide adequate precision and variance around the mean effect^15^. Combining acute, subacute, and chronic injury phases limits our ability to determine optimal timing of aerobic exercise delivery; future studies should examine these phases separately. Inter-trainer variability in rapport and instruction may have introduced inconsistencies. The unbalanced sex distribution in the aerobic group precluded sex-difference analyses; future studies should balance groups to test sex-specific effects. Additionally, while our three cognitive tests represent standard clinical markers, a more comprehensive battery may capture additional cognitive changes. Finally, the interpretation of the PROMIS Global Physical Health result warrants caution, as baseline differences between groups may have contributed to regression to the mean.

## Conclusion

A virtual, supervised, and individualized exercise intervention is both feasible and acceptable in community-based individuals with mTBI. Supervised aerobic exercise is potentially effective at improving executive function deficits observed post-mTBI. A larger randomized controlled trial is necessary to definitively assess the efficacy of the study.

## Data Availability

All data produced in the present study are available upon reasonable request to the authors

## Acknowledgements

We would like to thank the participants for their participation in this study. We would also like to thank the TBI Lab undergraduate research assistants for their help in administering and monitoring the interventions. Additionally, this work was done in part with the Discovery and Explorer Cluster, supported by Northeastern University’s Research Computing team. The data was partially collected using REDCap, supported by NIH UL1TR002544.

Data will be available upon request and uploaded to fitbir repository.

Study was funded by Northeastern University, specifically TPM start-up funding and the Institute for Cognitive and Brain Health Fellowship (ET).

## Bibliography

1. Maas AIR, Menon DK, Manley GT, et al. Traumatic brain injury: progress and challenges in prevention, clinical care, and research. Lancet Neurol. 2022;21(11):1004–1060. doi:10.1016/S1474-4422(22)00309-X

2. Hiskens MI. Targets of Neuroprotection and Review of Pharmacological Interventions in Traumatic Brain Injury. J Pharmacol Exp Ther. 2022;382(2):149–166. doi:10.1124/jpet.121.001023

3. Cabral DF, Rice J, Morris TP, Rundek T, Pascual-Leone A, Gomes-Osman J. Exercise for Brain Health: An Investigation into the Underlying Mechanisms Guided by Dose. Neurotherapeutics. 2019;16(3):580–599. doi:10.1007/s13311-019-00749-w

4. Alarie C, Gagnon I, Quilico E, Teel E, Swaine B. Physical Activity Interventions for Individuals With a Mild Traumatic Brain Injury:: A Scoping Review. J Head Trauma Rehabil. 2021;36(3):205. doi:10.1097/HTR.0000000000000639

5. Morris T, Gomes Osman J, Tormos Muñoz JM, Costa Miserachs D, Pascual Leone A. The role of physical exercise in cognitive recovery after traumatic brain injury: A systematic review. Restor Neurol Neurosci. 2016;34(6):977–988. doi:10.3233/RNN-160687

6. Pawlowski J, Dixon-Ibarra A, Driver S. Review of the Status of Physical Activity Research for Individuals With Traumatic Brain Injury. Arch Phys Med Rehabil. 2013;94(6):1184–1189. doi:10.1016/j.apmr.2013.01.005

7. Sharma B, Allison D, Tucker P, Mabbott D, Timmons BW. Cognitive and neural effects of exercise following traumatic brain injury: A systematic review of randomized and controlled clinical trials. Brain Inj. 2020;34(2):149–159. doi:10.1080/02699052.2019.1683892

8. Snowden T, Morrison J, Boerstra M, et al. Brain changes: aerobic exercise for traumatic brain injury rehabilitation. Front Hum Neurosci. 2023;17. doi:10.3389/fnhum.2023.1307507

9. Larson-Dupuis C, Léveillé E, Desjardins M, et al. Subtle long-term cognitive effects of a single mild traumatic brain injury and the impact of a three-month aerobic exercise intervention. J Sports Med Phys Fitness. 2021;61(1):87–95. doi:10.23736/S0022-4707.20.10918-6

10. Snyder AR, Greif SM, Clugston JR, et al. The Effect of Aerobic Exercise on Concussion Recovery: A Pilot Clinical Trial. J Int Neuropsychol Soc. 2021;27(8):790–804. doi:10.1017/S1355617721000886

11. Ding K, Tarumi T, Tomoto T, et al. A proof-of-concept trial of a community-based aerobic exercise program for individuals with traumatic brain injury. Brain Inj. 2021;35(2):233–240. doi:10.1080/02699052.2020.1865569

12. Chin LM, Keyser RE, Dsurney J, Chan L. Improved Cognitive Performance Following Aerobic Exercise Training in People with Traumatic Brain Injury. Arch Phys Med Rehabil. 2015;96(4):754. doi:10.1016/J.APMR.2014.11.009

13. Driver S, Ede A, Dodd Z, Stevens L, Warren AM. What barriers to physical activity do individuals with a recent brain injury face? Disabil Health J. 2012;5(2):117–125. doi:10.1016/j.dhjo.2011.11.002

14. Leddy JJ, Willer B. Use of Graded Exercise Testing in Concussion and Return-to-Activity Management. Curr Sports Med Rep. 2013;12(6):370. doi:10.1249/JSR.0000000000000008

15. Julious SA. Sample size of 12 per group rule of thumb for a pilot study. Pharm Stat. 2005;4(4):287–291. doi:10.1002/pst.185

16. Teresi JA, Yu X, Stewart AL, Hays RD. Guidelines for Designing and Evaluating Feasibility Pilot Studies. Med Care. 2022;60(1):95. doi:10.1097/MLR.0000000000001664

17. Tinney EM, Nwakamma MC, España-Irla G, et al. The Exercise and Concussion Health Study (TECHS): Pilot and Feasibility Protocol. Contemp Clin Trials Commun. Published online January 29, 2026:101608. doi:10.1016/j.conctc.2026.101608

18. Corrigan JD, Bogner J. Initial reliability and validity of the Ohio State University TBI Identification Method. J Head Trauma Rehabil. 2007;22(6):318–329. doi:10.1097/01.HTR.0000300227.67748.77

19. Bowie CR, Harvey PD. Administration and interpretation of the Trail Making Test. Nat Protoc. 2006;1(5):2277–2281. doi:10.1038/nprot.2006.390

20. Henry JD, Crawford JR. A Meta-Analytic Review of Verbal Fluency Performance in Patients With Traumatic Brain Injury. Neuropsychology. 2004;18(4):621–628. doi:10.1037/0894-4105.18.4.621

21. Brandt J. The hopkins verbal learning test: Development of a new memory test with six equivalent forms. Clin Neuropsychol. 1991;5(2):125–142. doi:10.1080/13854049108403297

22. Miutz LN, Burma JS, Brassard P, Phillips AA, Emery CA, Smirl JD. Comparison of the Buffalo Concussion Treadmill Test With a Physiologically Informed Cycle Test: Calgary Concussion Cycle Test. Sports Health. 2024;16(5):837–850. doi:10.1177/19417381231217744

23. Astrand PO, Ryhming I. A nomogram for calculation of aerobic capacity (physical fitness) from pulse rate during sub-maximal work. J Appl Physiol. 1954;7(2):218–221. doi:10.1152/jappl.1954.7.2.218

24. Borg G. Borg’s Perceived Exertion and Pain Scales. Human Kinetics; 1998:viii, 104.

25. Bell DR, Guskiewicz KM, Clark MA, Padua DA. Systematic Review of the Balance Error Scoring System. Sports Health. 2011;3(3):287–295. doi:10.1177/1941738111403122

26. Buysse DJ, Reynolds CF, Monk TH, Berman SR, Kupfer DJ. The Pittsburgh Sleep Quality Index: a new instrument for psychiatric practice and research. Psychiatry Res. 1989;28(2):193–213. doi:10.1016/0165-1781(89)90047-4

27. Cella D, Riley W, Stone A, et al. The Patient-Reported Outcomes Measurement Information System (PROMIS) developed and tested its first wave of adult self-reported health outcome item banks: 2005–2008. J Clin Epidemiol. 2010;63(11):1179–1194. doi:10.1016/j.jclinepi.2010.04.011

28. Cella D, Yount S, Rothrock N, et al. The Patient-Reported Outcomes Measurement Information System (PROMIS): Progress of an NIH Roadmap Cooperative Group During its First Two Years. Med Care. 2007;45(5):S3. doi:10.1097/01.mlr.0000258615.42478.55

29. Cella D, Choi SW, Condon DM, et al. PROMIS® Adult Health Profiles: Efficient Short-Form Measures of Seven Health Domains. Value Health J Int Soc Pharmacoeconomics Outcomes Res. 2019;22(5):537–544. doi:10.1016/j.jval.2019.02.004

30. Bassett DR. International physical activity questionnaire: 12-country reliability and validity. Med Sci Sports Exerc. 2003;35(8):1396. doi:10.1249/01.MSS.0000078923.96621.1D

31. Stubbs B, Vancampfort D, Rosenbaum S, et al. Dropout from exercise randomized controlled trials among people with depression: A meta-analysis and meta regression. J Affect Disord. 2016;190:457–466. doi:10.1016/j.jad.2015.10.019

32. Vancampfort D, Van Damme T, Brunner E, et al. Dropout From Exercise Interventions in Adults With Fibromyalgia: A Systematic Review and Meta-analysis. Arch Phys Med Rehabil. 2024;105(3):571–579. doi:10.1016/j.apmr.2023.06.002

33. López LP, Coll-Andreu M, Torras-Garcia M, et al. Aerobic exercise and cognitive function in chronic severe traumatic brain injury survivors: a within-subject A-B-A intervention study. BMC Sports Sci Med Rehabil. 2024;16(1):201. doi:10.1186/s13102-024-00993-4

34. Lange RT, Iverson GL, Zakrzewski MJ, Ethel-King PE, Franzen MD. Interpreting the Trail Making Test Following Traumatic Brain Injury: Comparison of Traditional Time Scores and Derived Indices. J Clin Exp Neuropsychol. 2005;27(7):897–906. doi:10.1080/1380339049091290

35. Iverson GL, Lange RT, Green P, Franzen MD. Detecting Exaggeration and Malingering With the Trail Making Test. Clin Neuropsychol. 2002;16(3):398–406. doi:10.1076/clin.16.3.398.13861

36. Ross SR, Millis SR, Rosenthal M. Neuropsychological prediction of psychosocial outcome after traumatic brain injury. Appl Neuropsychol. 1997;4(3):165–170. doi:10.1207/s15324826an0403_4

37. Ip RY, Dornan J, Schentag C. Traumatic brain injury: Factors predicting return to work or school. Brain Inj. 1995;9(5):517–532. doi:10.3109/02699059509008211

38. Heyder K, Suchan B, Daum I. Cortico-subcortical contributions to executive control. Acta Psychol (Amst). 2004;115(2):271–289. doi:10.1016/j.actpsy.2003.12.010

39. Tinney EM, Ai M, España-Irla G, Hillman CH, Morris TP. Physical activity and frontoparietal network connectivity in traumatic brain injury. Brain Behav. 2024;14(9):e70022. doi:10.1002/brb3.70022

40. España-Irla G, Tinney EM, Ai M, Nwakamma M, Morris TP. Functional Connectivity Patterns Following Mild Traumatic Brain Injury and the Association With Longitudinal Cognitive Function. Hum Brain Mapp. 2025;46(8):e70237. doi:10.1002/hbm.70237

41. Rogge AK, Röder B, Zech A, et al. Balance training improves memory and spatial cognition in healthy adults. Sci Rep. 2017;7(1):5661. doi:10.1038/s41598-017-06071-9

42. Schiff ND, Giacino JT, Butson CR, et al. Thalamic deep brain stimulation in traumatic brain injury: a phase 1, randomized feasibility study. Nat Med. 2023;29(12):12. doi:10.1038/s41591-023-02638-4

43. Mathias JL, Alvaro PK. Prevalence of sleep disturbances, disorders, and problems following traumatic brain injury: A meta-analysis. Sleep Med. 2012;13(7):898–905. doi:10.1016/j.sleep.2012.04.006

44. Tinney EM, España-Irla G, Warren AEL, et al. Axonal injury, sleep disturbances, and memory following traumatic brain injury. Ann Clin Transl Neurol. n/a(n/a). doi:10.1002/acn3.52145

45. Fiorin F da S, Godinho DB, dos Santos EB, et al. Relationship among depression, fatigue, and sleep after traumatic brain injury: The role of physical exercise as a non-pharmacological therapy. Exp Neurol. 2025;386:115156. doi:10.1016/j.expneurol.2025.115156

46. Korkutata A, Korkutata M, Lazarus M. The impact of exercise on sleep and sleep disorders. Npj Biol Timing Sleep. 2025;2(1):5. doi:10.1038/s44323-024-00018-w

47. Damiano DL, Zampieri C, Ge J, Acevedo A, Dsurney J. Effects of a rapid-resisted elliptical training program on motor, cognitive and neurobehavioral functioning in adults with chronic traumatic brain injury. Exp Brain Res. 2016;234(8):2245–2252. doi:10.1007/s00221-016-4630-8

48. Terwee CB, Peipert JD, Chapman R, et al. Minimal important change (MIC): a conceptual clarification and systematic review of MIC estimates of PROMIS measures. Qual Life Res Int J Qual Life Asp Treat Care Rehabil. 2021;30(10):2729–2754. doi:10.1007/s11136-021-02925-y

49. Morris TP, Fried PJ, Macone J, et al. Light aerobic exercise modulates executive function and cortical excitability. Eur J Neurosci. 2020;51(7):1723–1734. doi:10.1111/ejn.14593

50. Mansoor M, Ibrahim A, Hamide A, Tran T, Candreva E, Baltaji J. Exercise-Induced Neuroplasticity: Adaptive Mechanisms and Preventive Potential in Neurodegenerative Disorders. Physiologia. 2025;5(2):13. doi:10.3390/physiologia5020013

