## Supplementary Material for "The feasibility and efficacy of a virtual, symptom-guided aerobic exercise intervention to improve cognition in mild traumatic brain injury: A single-blind pilot randomized control trial with an active comparator group"

Supplementary Materials

Supplementary Material 1: Statistical Approaches

Post-Intervention Group Comparison. We conducted Analysis of Covariance (ANCOVA) to compare post-intervention scores between groups while controlling for baseline performance:

*Post-Score ~ Group + Pre-Score + Age + Sex + Education + Days Since Injury + [Selected Covariates].*

The *Group* coefficient represented the adjusted mean difference in post-intervention outcomes between treatments. ANCOVA assumptions were checked using normality of residuals (Shapiro-Wilk test), homogeneity of variance (Levene's test), and homogeneity of regression slopes (Group × Pre-Score interaction test). Outliers were identified using standardized residuals (|r| > 3) and Cook's distance (D > 4/n), but not removed due to the nature of the pilot study. See supplementary materials for graphs of all assumptions. We calculated Hedges' g to quantify effect sizes and account for sample size. For each outcome, we reported both a base model (fixed covariates only) and a final model (with selected optional covariates), along with the AIC improvement.

Group × Time Interaction. We fitted linear mixed-effects models with the formula:

*Score ~ Group × Time + Age + Sex + Education + Days Since Injury + [Selected Covariates] + (1|Participant),*

where *Group* was treatment assignment (aerobic vs. balance), *Time* was assessment timepoint (pre vs. post), and a random intercept accounted for within-person correlation. The *Group × Time* interaction coefficient quantified differential change between groups from pre- to post-intervention. The models were fitted using restricted maximum likelihood estimation with the *lme4* package in R. We calculated Hedges' g for the interaction effect. We report base and final model results with AIC improvement.

These two approaches addressed complementary questions: The ANCOVA compared end-point outcomes after adjusting for baseline (post-intervention differences). The group x time interaction tests whether the rate of change differs between groups (differential trajectories). Effect sizes were interpreted using conventional benchmarks: |g| < 0.2 = negligible, 0.2 ≤ |g| < 0.5 = small, 0.5 ≤ |g| < 0.8 = medium, |g| ≥ 0.8 = large. All analyses were conducted in R using *lme4*, *lmerTest*, *car*, *effectsize*, and *broom* packages.

Supplementary Material 2: Assumptions

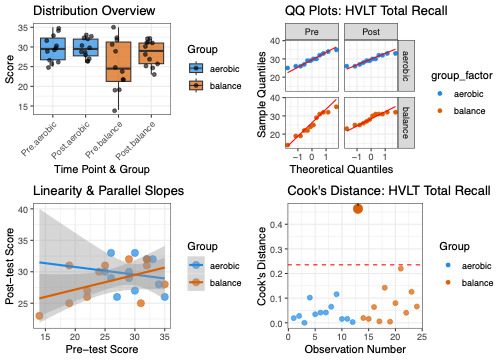

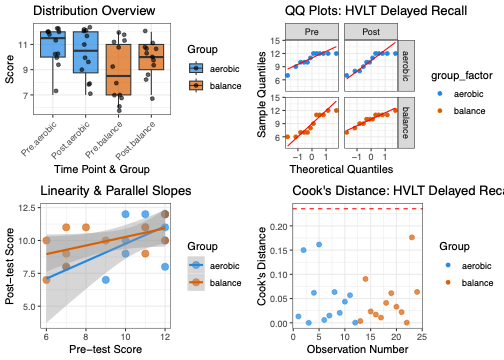

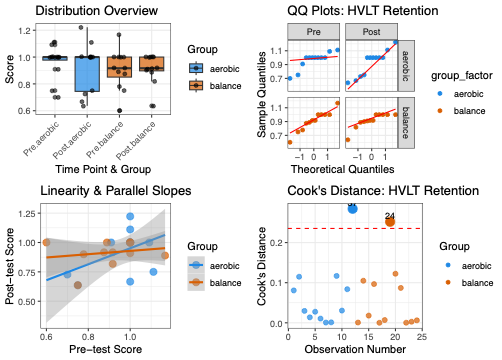

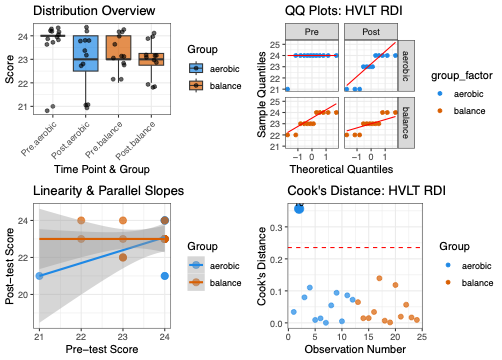

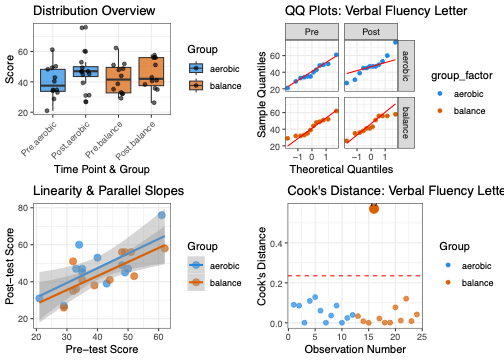

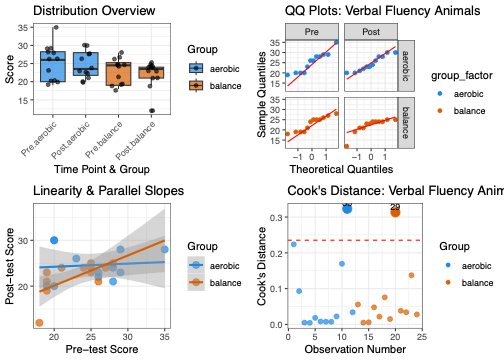

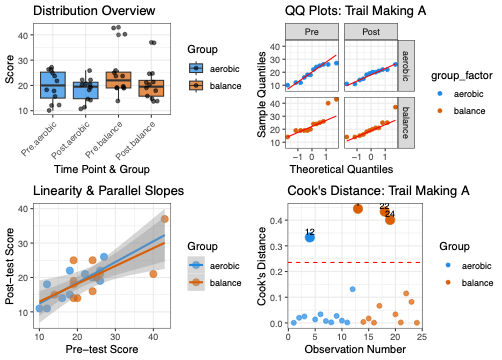

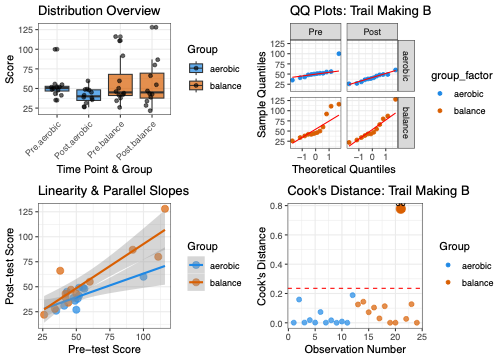

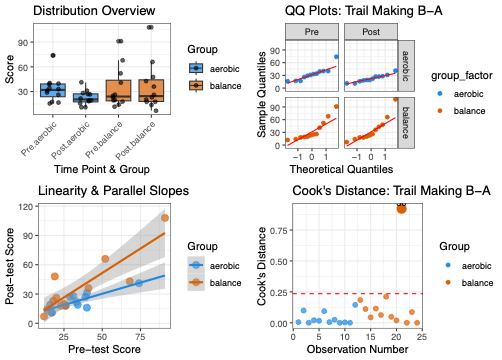

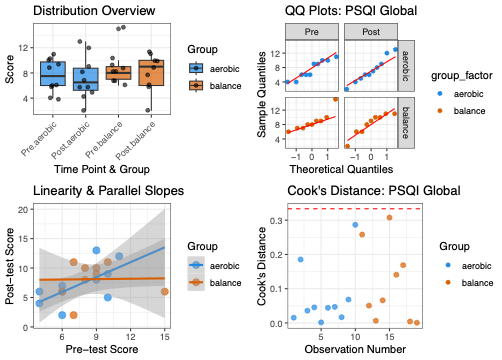

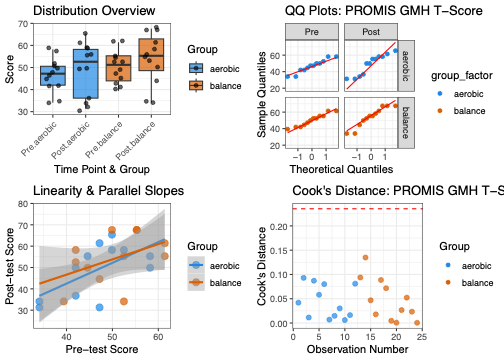

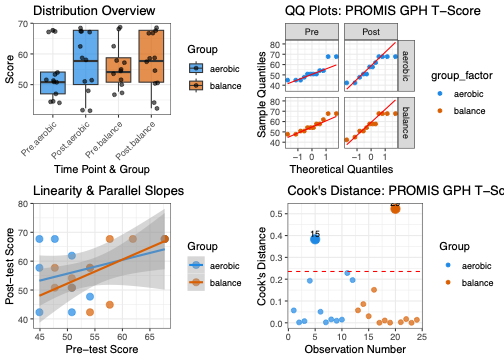

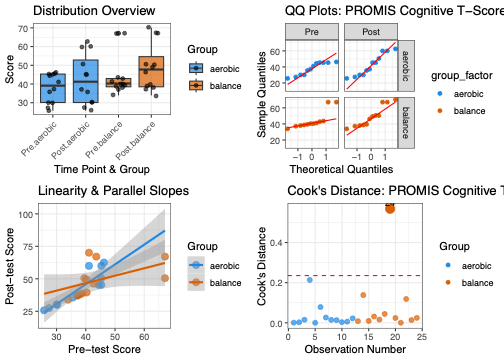

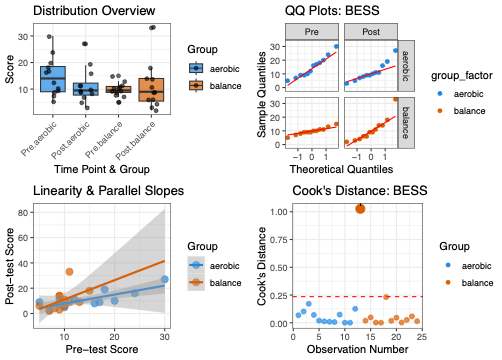

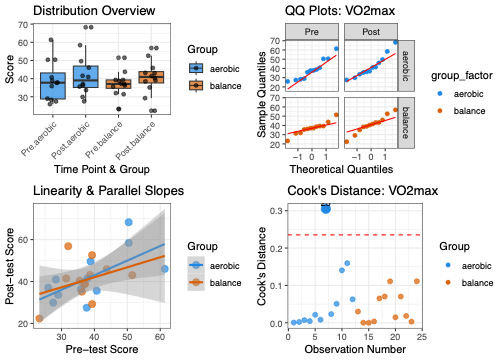

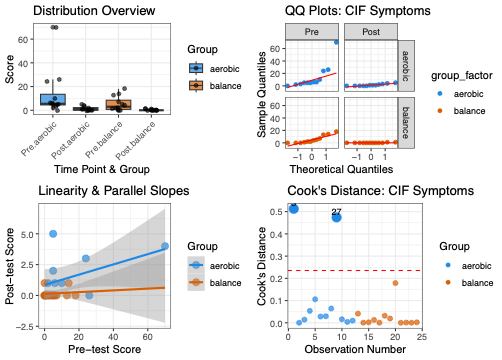

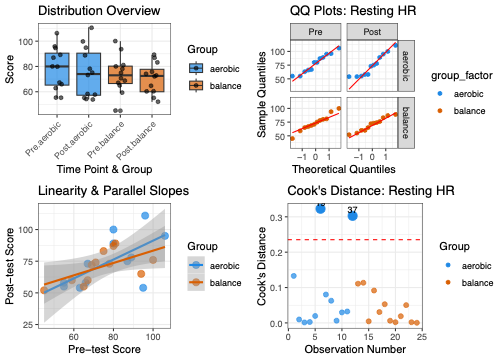

Supplementary Material 3 – Concussion like symptoms and severity at baseline

| **Symptoms** | **# of participants with symptoms** | **Total symptom severity** |
| --- | --- | --- |
| Nausea | 4 | 5 |
| Headache | 13 | 21 |
| Pressure | 8 | 14 |
| Neck | 6 | 12 |
| Dizzy vomit | 6 | 9 |
| Vision | 2 | 4 |
| Balance | 7 | 11 |
| Light sensitivity | 15 | 25 |
| Noise sensitivity | 5 | 9 |
| Slow | 8 | 23 |
| Fog | 13 | 30 |
| Feel right | 6 | 15 |
| Concentration | 9 | 25 |
| Remember | 11 | 25 |
| Fatigue | 14 | 32 |
| Confusion | 3 | 6 |
| Drowsiness | 5 | 10 |
| Sleep | 3 | 11 |
| Emotional | 4 | 16 |
| Irritability | 7 | 21 |
| Sad | 3 | 12 |
| Anxious | 9 | 23 |

Table 1: Table displaying the total number of participants reporting concussion-like symptoms at baseline (prior to beginning the MBET) and total severity of report symptoms.

Supplementary Material 4: All Graphs

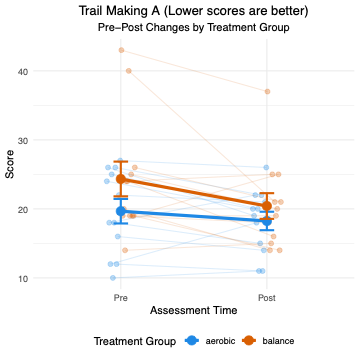

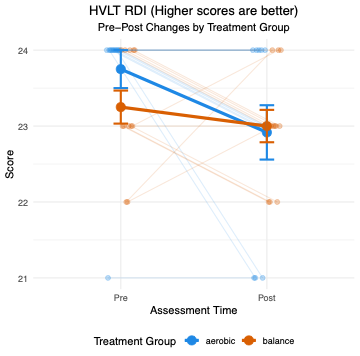

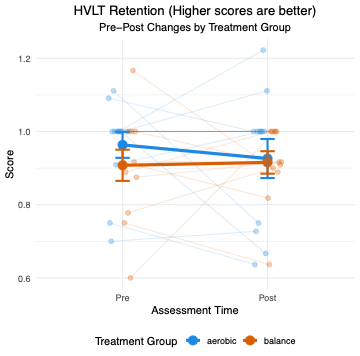

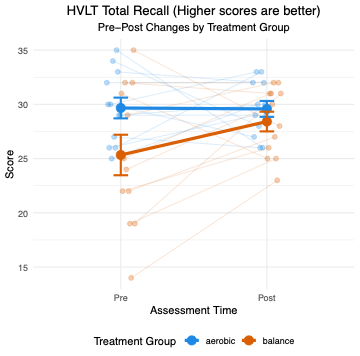

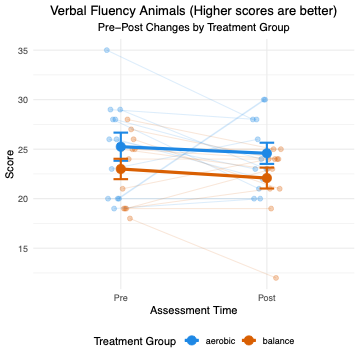

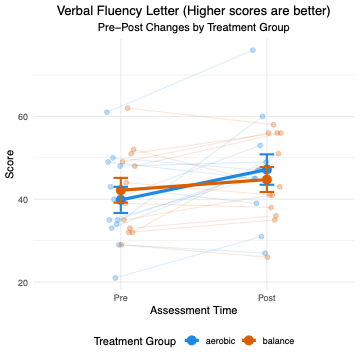

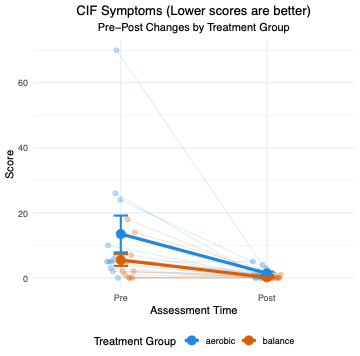

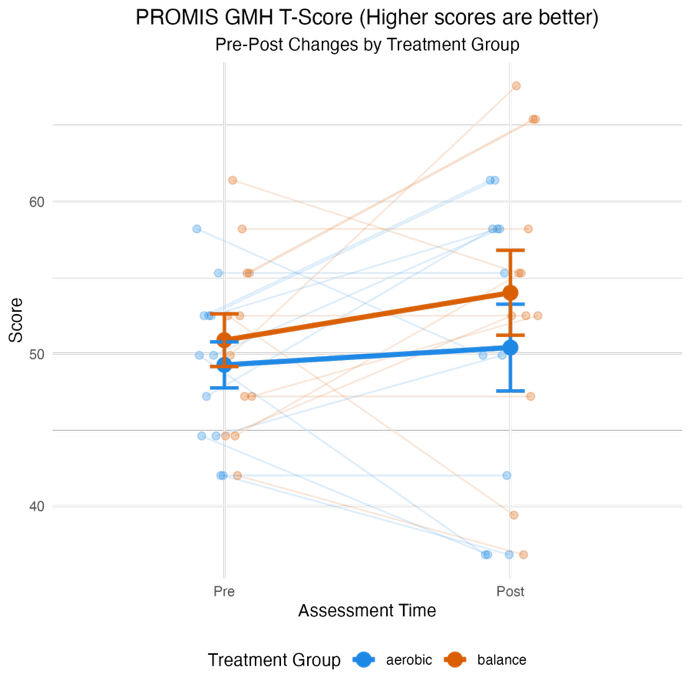

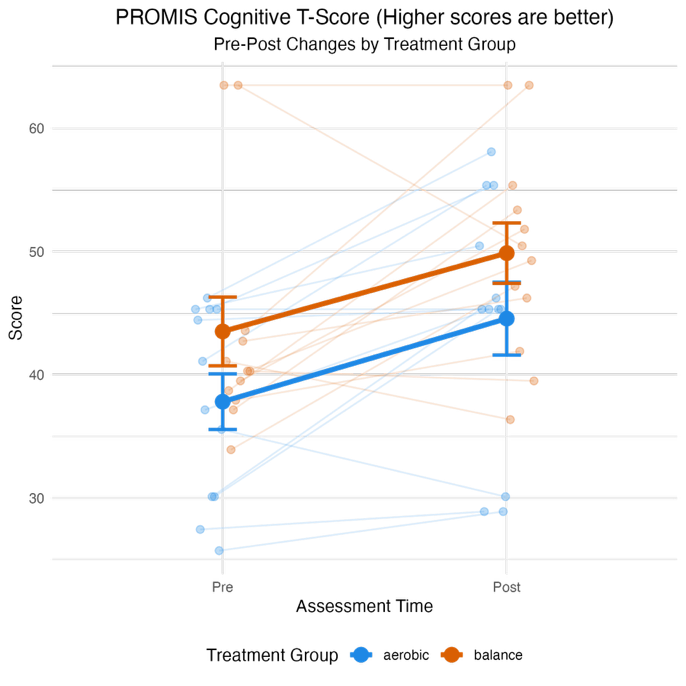

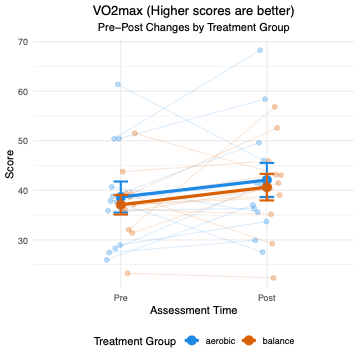

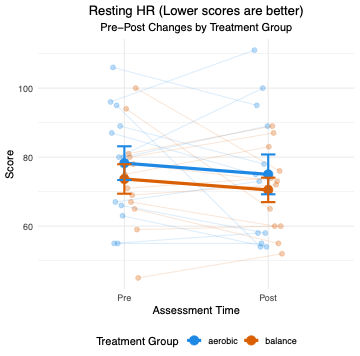

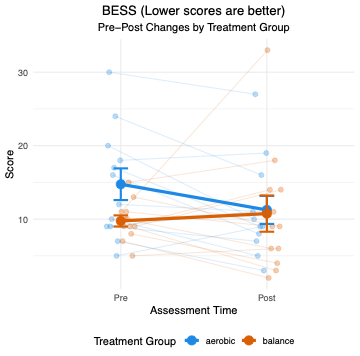

Supplementary Material 5: Correlation with time in HR zone and outcome change scores

To test if there were associations time spent in HR zone and change in outcome measure, we performed a t-test using change scores for TMT-b, TMT b-a, HVLT DR, and PSQI. We report the correlation coefficient (r) and 95% CI and correlation graphs.

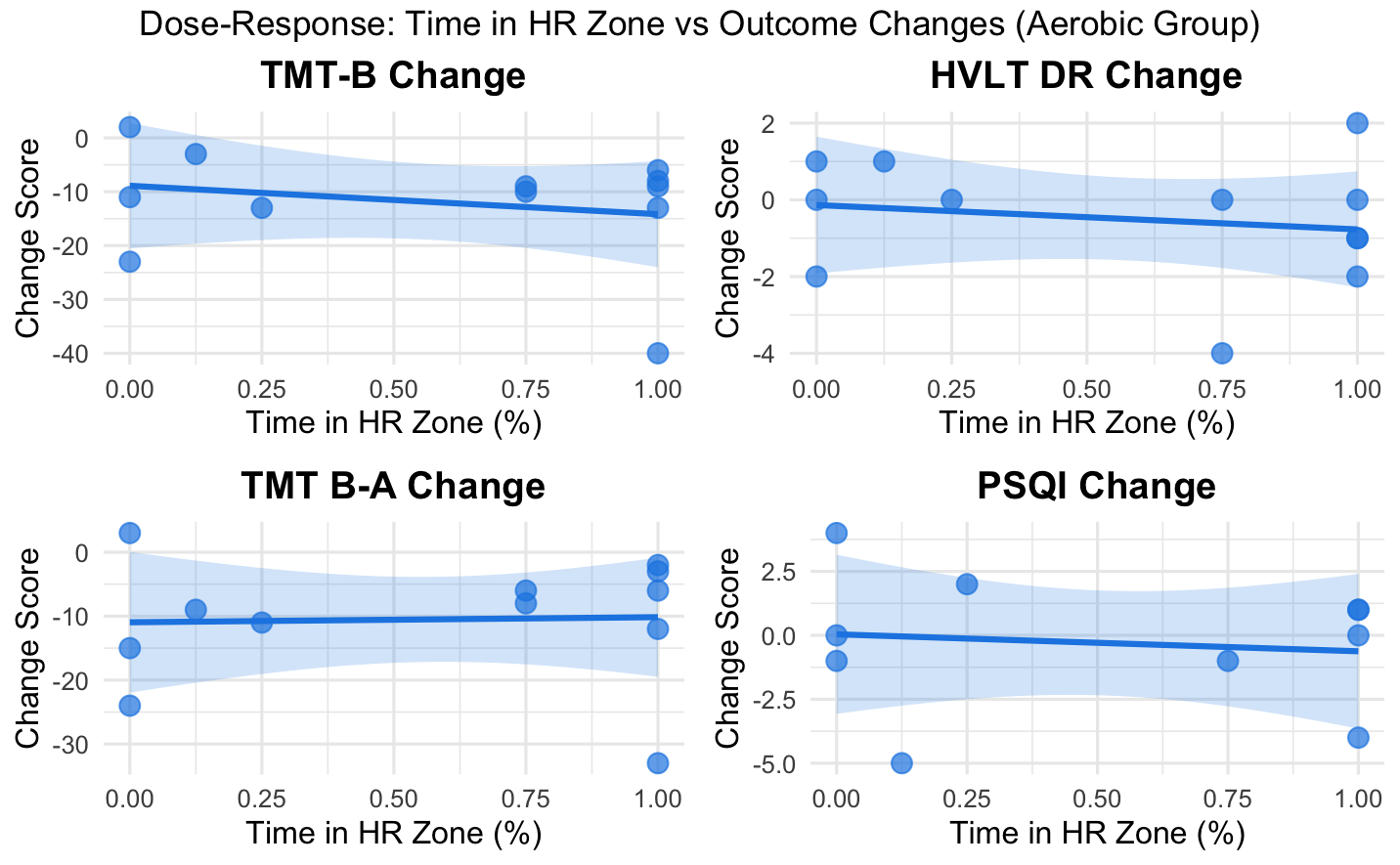

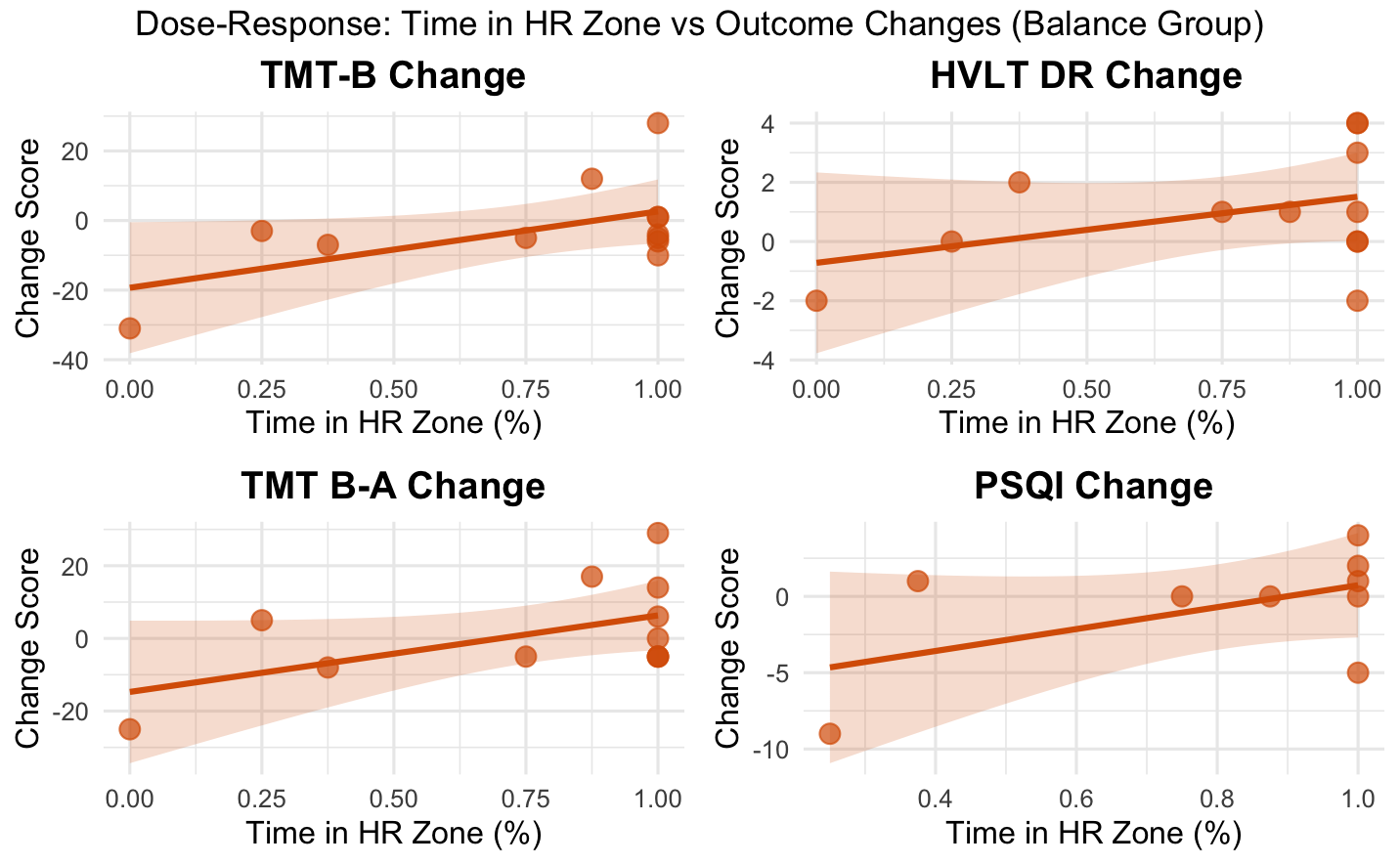

| Aerobic Correlation (r, [95% CI]) | |
| --- | --- |
| TMT B | (-0.224, [-0.71, 0.40]) |
| TMT B-A | (0.04, [-0.55, 0.59]) |
| HVLT DR | (-0.177, [-0.68, 0.44]) |
| PSQI | (-0.12, [-0.69, 0.55]) |
| Balance Correlation (r, [95% CI]) | |
| TMT B | (0.57, [-0.01, 0.86]) |
| TMT B-A | (0.54, [-0.05, 0.85]) |
| HVLT DR | (0.39, [-0.23, 0.79]) |
| PSQI | (0.54, [-0.19, 0.89]) |

Supplementary Material 6: Does Number of Prior Injuries change Treatment Response?

To test if there were differences between 0-1 prior injuries and 2+ prior injuries, we performed a t-test using change scores for TMT-b, TMT b-a, HVLT DR, and PSQI. We report Cohen’s d and 95% CI and show box plots.

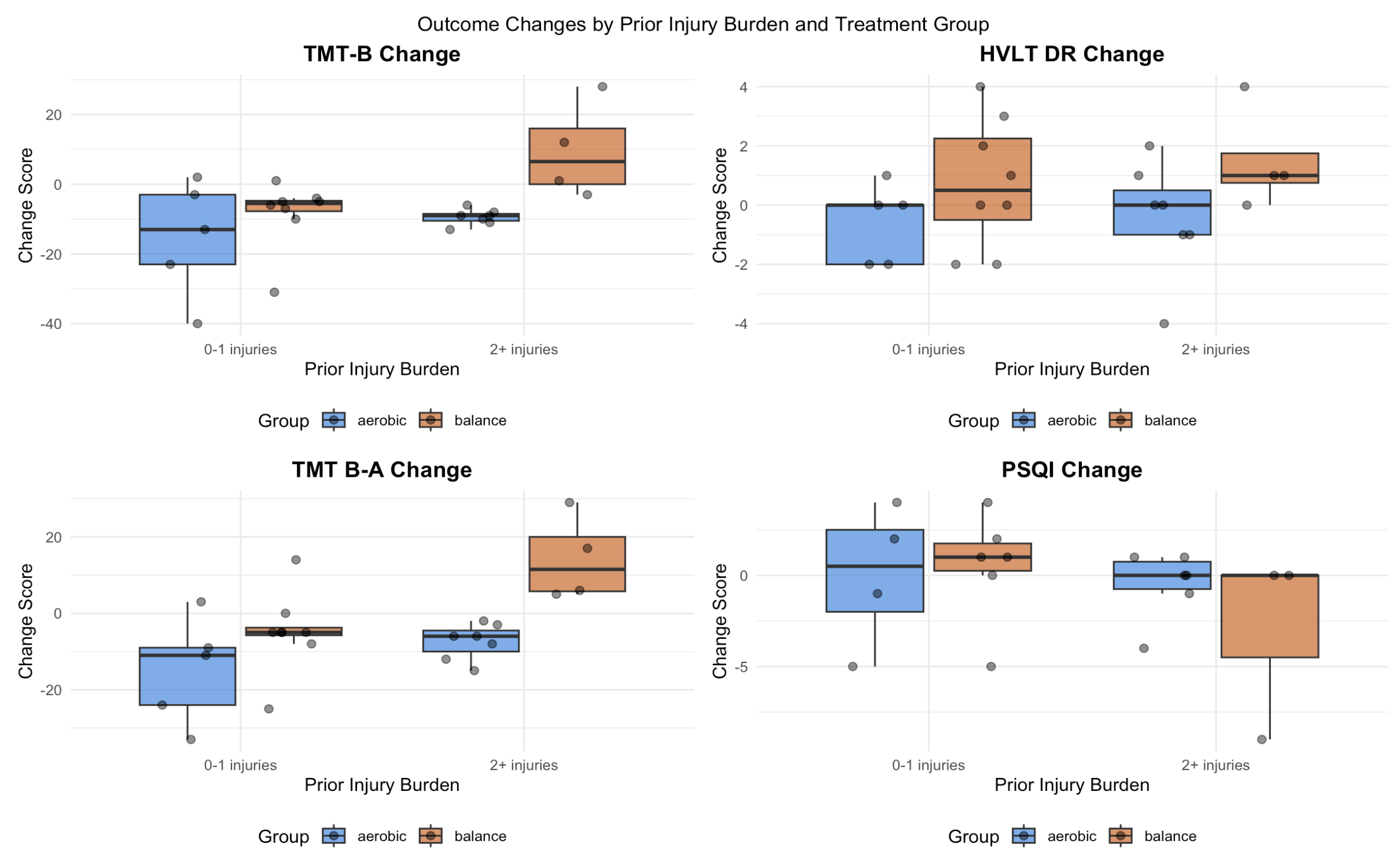

| Aerobic Difference (d, [95% CI]) | |
| --- | --- |
| TMT B | 0.56 [-14.78, 26.73] |
| TMT B-A | 0.77 [-9.71, 24.45] |
| HVLT DR | 0.10 [-1.92, 2.26] |
| PSQI | -0.18 [-6.38, 5.38] |
| Balance Difference (d, [95% CI]) | |
| TMT B | 1.61 [-2.65, 38.40] |
| TMT B-A | 0.36 [-1.95, 3.45] |
| HVLT DR | 1.77 [2.44, 35.81] |
| PSQI | -0.93 [-14.50, 7.50] |

Supplementary Material 7: Correlation with Adherence and outcome change scores

To test if there were associations adherence rate and change in outcome measure, we performed a t-test using change scores for TMT-b, TMT b-a, HVLT DR, and PSQI. We report the correlation coefficient (r) and 95% CI and correlation graphs.

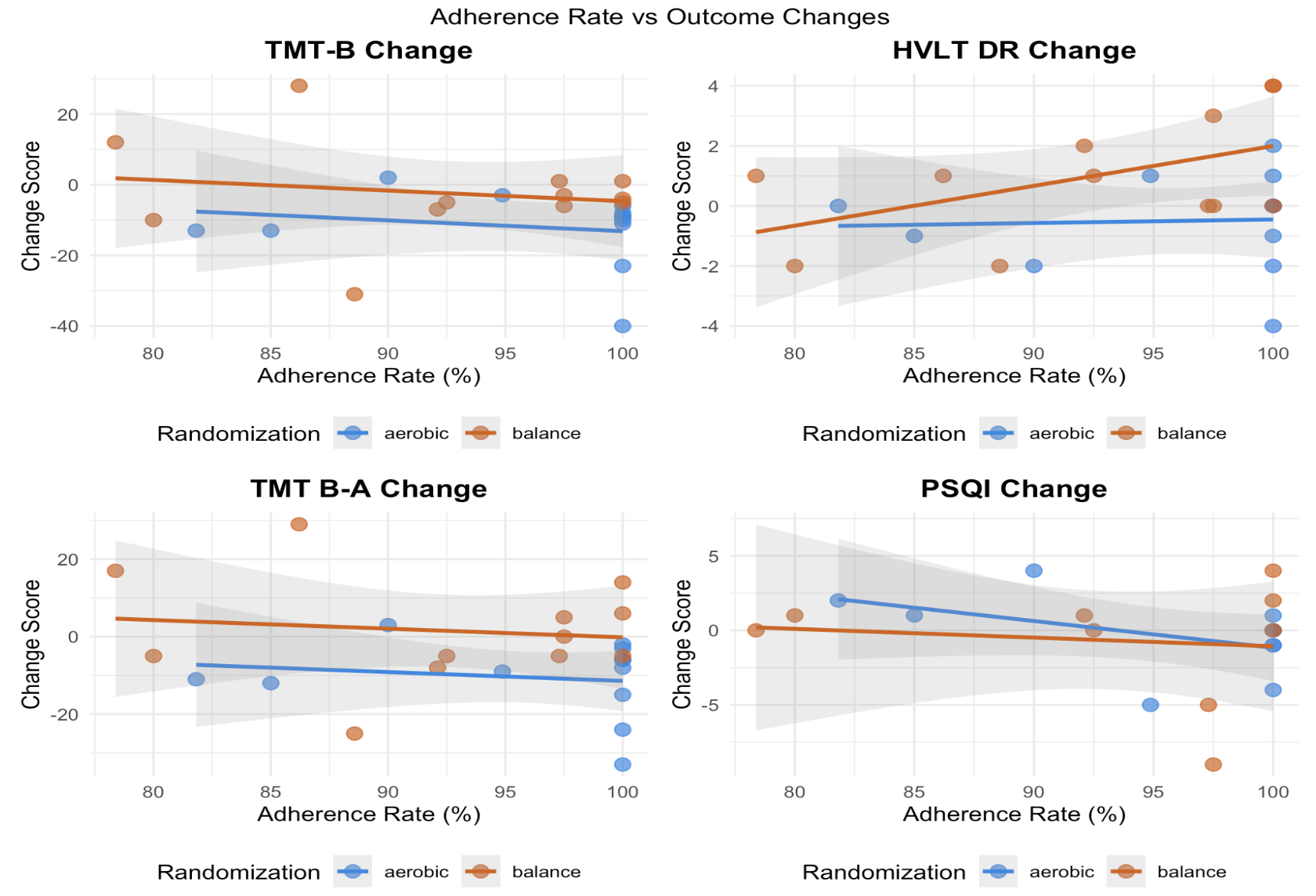

| Aerobic Correlation (r, [95% CI]) | |
| --- | --- |
| TMT B | (-0.19, [-0.69, 0.43]) |
| TMT B-A | (-0.15, [-0.54, 0.61]) |
| HVLT DR | (0.05, [-0.09, 0.84]) |
| PSQI | (-0.47, [-0.85, 0.22]) |
| Balance Correlation (r, [95% CI]) | |
| TMT B | (-0.17, [-0.68, 0.45]) |
| TMT B-A | (-0.12, [-0.65, 0.49]) |
| HVLT DR | (0.51, [-0.09, 0.84]) |
| PSQI | (-0.13, [-0.73, 0.59]) |

Supplementary Material 8: AIC Model Building

By using AIC model building, we were able to better fit our models using specific covariates, providing valuable insights for developing personalized treatment approaches. Specifically, time in HR zone, adherence rate, and number of prior injuries were consistent in improving model fit across multiple measures. No correlations emerged between time in HR zone and cognitive or sleep improvements, suggesting that exceeding the prescribed intensity threshold may not confer additional cognitive or sleep benefits. The number of prior injuries altered treatment response for the balance group in delayed recall; however, samples were extremely small in each group, so this should be interpreted with caution. Although, if the number of prior injuries does determine treatment response, that has large implications for precision medicine approaches, and supports the notion that injury history should be considered when prescribing exercise intensity and duration. Within the aerobic group, adherence was not associated with improvements, while in the balance group, adherence showed stronger influence on outcomes. This may be due to our higher adherence rate in the aerobic group compared to balance. The moderation analyses revealed that adherence did not moderate group differences, suggesting that both interventions benefited similarly from high adherence.

Supplementary Material 9: Feedback Satisfaction Survey Results

| Question | Mean (SD) |
| --- | --- |
| Do you believe your participation in this study will contribute to valuable and meaningful patient outcomes? | 96% yes |
| On a scale of 1 (very dissatisfied) to 10 (extremely satisfied), how satisfied were you with the organization, clarity of the content, and communication between you and the lab members during the pre-screening sessions? | 9.15 (1.31) |
| On a scale of 1 (very dissatisfied) to 10 (extremely satisfied), how satisfied were you with the baseline sessions? (i.e. think in terms of content, length, and how clearly and thoroughly the material from this portion of the study was explained) | 8.84 (1.82) |
| On a scale of 1 (very dissatisfied) to 10 (extremely satisfied), how satisfied were you with the exercise interventions in this study? (i.e. scheduling, communication, feasibility of the sessions, and how accommodating the exercise intervention leader was to your needs, etc.) | 8.61 (1.67) |
| On a scale of 1 (very dissatisfied) to 10 (very satisfied), how satisfied were you with the endpoint sessions of this study? (i.e. the length of the sessions and whether the content in this portion of the study was presented in a clear and thorough manner) | 9.34 (0.97) |
| On a scale of 1 (very dissatisfied) to 10 (very satisfied), how would you rate your overall experience with this study? | 8.57 (2.08) |
| Would you recommend participating in this study to a friend or family member? | 96% yes |
| Overall, how would you rate the communication between the you as a participant and the lab on a scale of 1 (very dissatisfied) to 10 (very satisfied)? | 8.80 (1.74) |
| Did you feel that you were adequately informed about how the study would proceed and what to expect as a participant? | 96% yes |
| Do you feel that your privacy and confidentiality were protected during this study? | 95% yes |
| On a scale of 1 (very dissatisfied) to 10 (very satisfied), how would you rate your experience with scheduling sessions in terms of flexibility and convenience? | 8.56 (1.73) |
| Was there anything you found extremely difficult while participating in this study? | - “The scheduling was a bit hard” - “I found that the exercise was exacerbating my concussion symptoms.” - “My ability to focus and maintain communication with the team was the most difficult part, and interfered with my ability to participate. This is on me, not the study team.” - “Overscheduling when I needed rest to recover from symptoms” - “Coordinating the exercise sessions was tricky sometimes but we always managed to figure it out. I have a very flexible schedule so that helped” - “Sometimes just finding the time for the sessions, but otherwise no” - “Wearing the monitor” - “Fitting in the exercise sessions with everyone's schedule was challenging, but I'm glad I did it.” |
| Is there anything you believe the lab could do to improve any part of the study for participants? | - “I think having a weekly scheduling portal would make the study flow a bit better.” - “When I was actively participating, I had expected to keep a regular schedule for my exercise. There was inconsistency and last minute changes. Keeping a consistent schedule would be an improvement.” - “The log/form that came with the actigraph watch was confusing” - “I think compensation should be higher given the time commitment it actually represents” - “Everything was executed to a high standard.” - “More consistency in scheduling and more communication about the timing of the watch would have been good. Also checking better on the battery life of items before they were given to me would have helped.” - “Same schedule and instructor each session” |
| At any point during the study, did you feel frustrated? | - “I feel that at some points, while I recognize that my participation in the study would potentially benefit individuals in the future, that it did not benefit me as much as I was anticipating. However, I feel that this is more so because I wasn't completely aware of what to expect from the interventions, and was placed in a more relaxed balance intervention as opposed to a more intense intervention.” - “A little by how tired I was with the intense workouts plus working a physical job and being in physical therapy at the same time” - “It was taking up a lot of time, and I wasn't seeing any obvious benefits of participating” - “Only as a concussion symptom!” - “When I was hospitalized the nurses wouldn't let me continue the zoom sessions with the study and that was when I was frustrated because I knew a break in communication would be difficult for me to overcome once I was discharged.” - “Yes. I had delayed onset concussion symptoms post exercise; sometimes did not feel up to exercising and skimped on needed rest to participate after a cognitively challenging day” - “Two times I had study items run out of batteries and once or twice they were late for the sessions, but overall I was very happy with everything” |
| Did you expect to receive any specific outcome(s) from participating in this study? | - “I was interested in seeing the results of the MRI but understand that’s not possible” - “I was hoping it would help speed my recovery, but I didn't experience that. - “I think participating in the study forced me to focus more on my health throughout each week, and I expected to become more in tune with my body. This was ultimately an outcome of the exercises I completed each week.” - “Better at balancing” - “I was hoping to have my concussion symptoms reduced, and I expect it would at least help me get in shape” - “Yes, improved fitness” - “I expected to learn more about my brain and how it works and I certainly did” - “Yes, improvement in symptoms which has been good” - “I was curious how my symptoms and condition would evolve” |
| Other comments | - “The TECHS study was an incredibly flexible, convenient, and easy study to participate in. As someone who has suffered from three concussions, I would strongly recommend other individuals to participate in this study as I feel that exercise can truly help improve the recovery period following a traumatic brain injury.” - “Participating in this study helped to alleviate some of my daily headaches as well as other concussion symptoms. Following the study, I've been able to implement exercises and what I learned about my own body during the study to continue the healing process. I find that on days where I have more symptoms, I'm more inclined to do a short period of exercise, and it often helps! I'm really grateful to have participated, and hope that the findings of this study can help other folks with TBI's as well.” - “Thanks, I was glad to participate and regretted that I couldn't continue” - “The structure of the study really helped me get my workouts back on track after having to take a lot of time off from exercising due to my concussion. I felt like I really didn't have the mental bandwidth to figure that out myself, because at the time I was really focused on getting back to working full time, it was exhausting. I sincerely appreciated the help and support in figuring out what exercise I could tolerate and in getting back on a more regular exercise schedule.” - “The study was an excellent experience! Everyone involved was highly professional, and the lab was lovely.” - “I was pleasantly surprised at how well the exercises improved my daily headaches” - “Staff was accommodating, flexible, I hope my participation in this study will be valuable to those who have suffered from concussions.” - “Thank you to all the staff and students who helped facilitate all my sessions! it was so lovely to engage with you all and I appreciate your hard work and commitment.” - “Towards the end of the study, my energy and activity levels improved greatly. Thanks to the intense focus on balance and strength training, I was able to resume my extreme sports. I felt assured that my body and mind had recovered. The trainers were fantastic! They consistently knew how to guide me and were incredibly supportive throughout all the training sessions.” - “Participating in research is the only way we can help others have an easier road than we have had. I strongly believe it is my duty to help providers learn from my experience for my children and all children. No one should have to walk TBI recovery without knowing what can help.” |

Supplementary Material 10: HR-symptom correlation over time in the aerobic exercise intervention
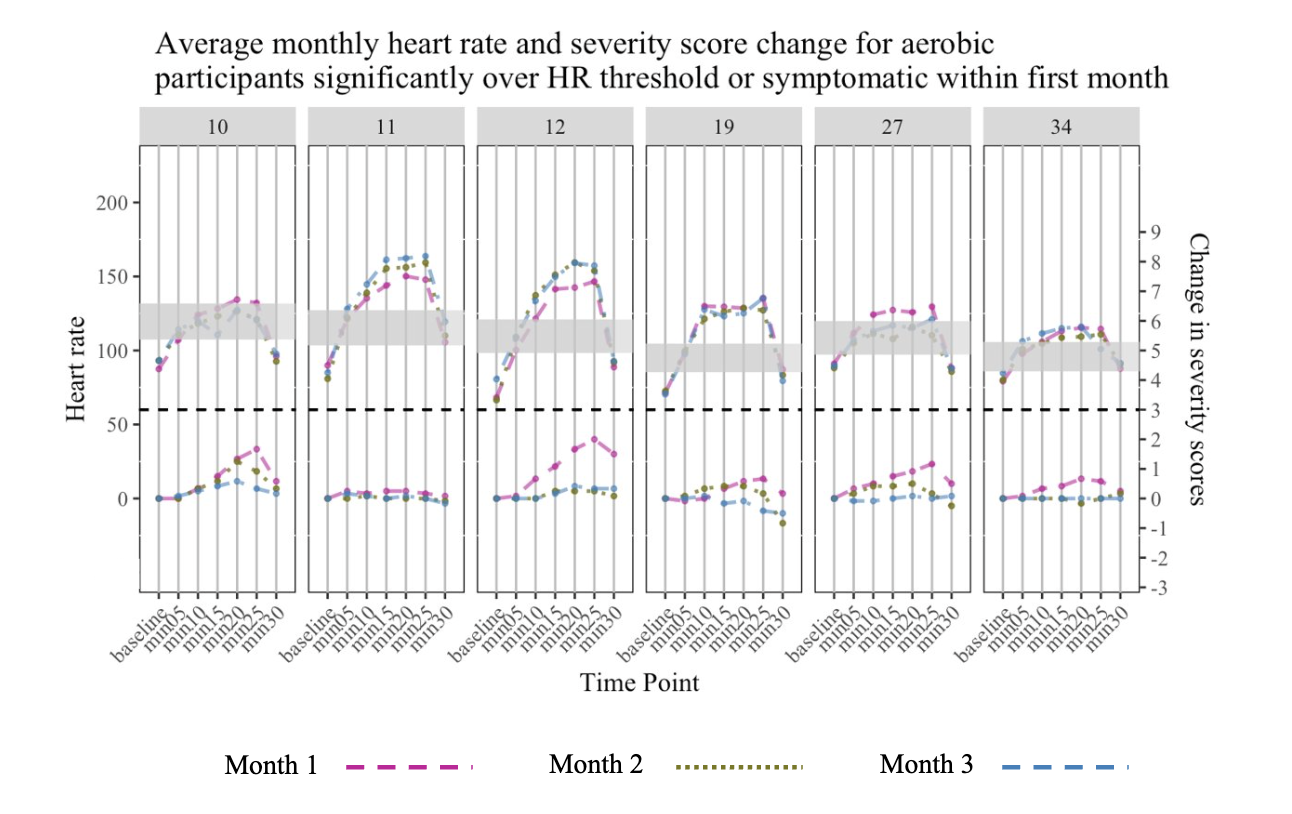

Six- representative participants, illustrating the decrease in HR-symptom correlation over time. Plots displaying participant-specific average change in HR (above black dotted line) and symptom severity (below black dotted line) over the 12-week (36 sessions) aerobic exercise intervention. The shaded gray area indicates the 65 – 80% of HR at symptom threshold/volitional exhaustion. The black dotted line indicates symptom threshold (three-unit exercise-induced symptom severity increase from baseline). The red dashed lines show changes in mean heart rate and symptom severity in the first 12 sessions (Month 1); the green dotted lines 13 to 24 (Month 2); and the blue dashed lines sessions 25 to 36 (Month 3).

Supplementary Material 11: Percent change in TMT-B in both groups. 10 out of 12 aerobic participants improved more than 10%, whereas only 5 out of 12 balance participants improved more than 10%. One participant did increase 73.68%, increasing 28 seconds from baseline (38 seconds) to endpoint (66 seconds). However, this participant was not identified as an influential outlier via Cook’s distance, and the raw change score is less than 3 standard deviations away from the mean.

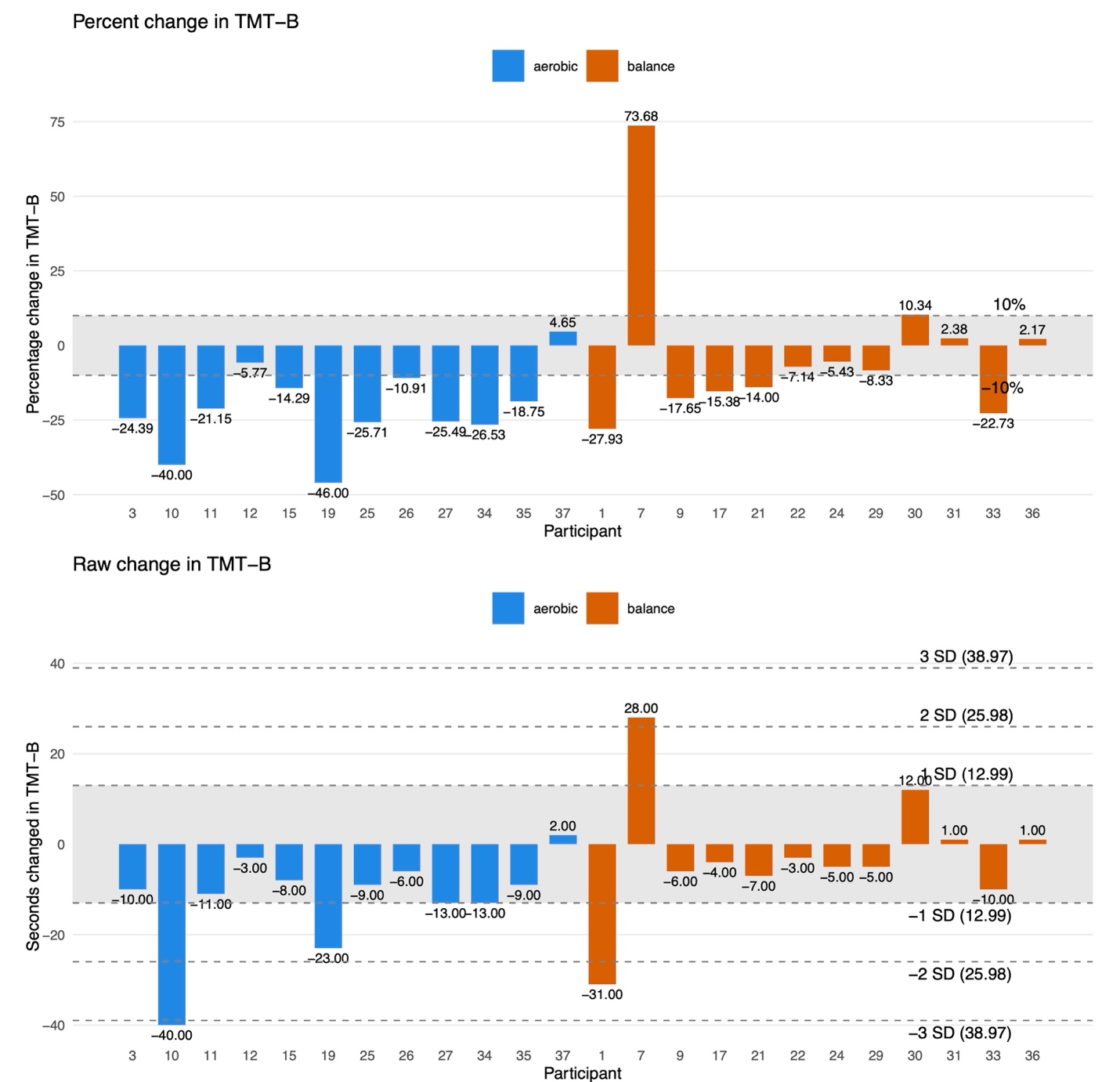

Percent change in TMT B test, with 10% change showing clinically significant improvement. Negative percentages show faster TMT B times at endpoint

Supplementary Material 12: Post intervention ANCOVA results

ANCOVA Post Intervention Differences

| **Measure** | **Base AIC** | **Base Hedges' g (95% CI)** | **Final AIC** | **AIC Change** | **β, SE** | **Final Hedges' g (95% CI)** | **Optional Covariates** |
| --- | --- | --- | --- | --- | --- | --- | --- |
| **Hopkin Verbal Learning Test Total Recall** | 129.65 | -0.24 [-1.47, 0.98] | 123.60 | 6.04 | 0.02, 1.66 | 0.01 [-1.25, 1.26] | Adherence Rate |
| **Hopkin Verbal Learning Test Delayed Recall** | 83.31 | 0.76 [-0.42, 1.94] | 81.24 | 2.07 | 1.44, 0.70 | 1.25 [-0.05, 2.55] | Adherence Rate, time in heart rate |
| **Hopkin Verbal Learning Test Retention** | -21.12 | 0.29 [-0.87, 1.45] | -26.82 | 5.69 | 0.08, 0.07 | 0.69 [-0.51, 1.89] | Baseline symptoms |
| **Hopkin Verbal Learning Test Recognition Discrimination Index** | 77.48 | 0.12 [-1.04, 1.28] | 71.09 | 6.39 | -0.05, 0.53 | -0.05 [-1.27, 1.17] | Number of prior injuries, Adherence Rate, total met |
| **Verbal Fluency Letter** | 173.82 | 0.03 [-1.13, 1.19] | 171.24 | 2.58 | 1.93, 4.24 | 0.26 [-0.93, 1.44] | Number of prior injuries |
| **Verbal Fluency Animals** | 130.00 | -1.29 [-2.57, -0.01] | 126.46 | 3.54 | -2.99, 1.89 | -1.01 [-2.35, 0.34] | Baseline VO_2_max |
| **Trail Making A** | 143.98 | -0.07 [-1.22, 1.09] | 141.38 | 2.6 | 1.15, 2.35 | 0.28 [-0.95, 1.51] | time in heart rate |
| **Trail Making B** | 191.28 | 0.86 [-0.31, 2.03] | 187.29 | 3.98 | 13.15, 6.05 | 1.24 [0.04, 2.45] | Number of prior injuries |
| **Trail Making B-A** | 192.58 | 0.87 [-0.30, 2.04] | 189.52 | 3.06 | 13.36, 6.31 | 1.20 [0.00, 2.41] | Number of prior injuries |
| **Pittsburg Sleep Quality Index Global** | 91.55 | 1.02 [-0.22, 2.26] | 89.75 | 1.79 | 3.75, 1.46 | 1.65 [0.22, 3.09] | Baseline symptoms, total met |
| **PROMIS Global Physical Health T-Score** | 164.61 | -0.724 [ -1.92, 0.473] | 162.75 | 4.32 | -7.97, 3.66 | -1.33 [-2.63, -0.03 ] | Number of prior injuries, time in heart rate |
| **PROMIS Global Mental Health T-Score** | 181.82 | -0.1 [-1.39, 1.19 ] | 180.98 | 0.84 | -0.81, 5.66 | -0.09 [-1.38, 1.21 ] | Number of prior injuries |
| **PROMIS Cognitive T-Score** | 176.72 | 0.459 [-0.699, 1.618 ] | 173.87 | 2.84 | 2.032, 4.496 | 0.254 [-0.937, 1.445 ] | Number of prior injuries |
| **Balance Error Scoring System** | 157.51 | -0.27 [-1.52, 0.98] | 157.51 | 0 | -1.57, 3.40 | -0.27 [-1.52, 0.98] | None |
| **Resting Heart Rate** | 204.50 | -0.04 [-1.22, 1.13] | 196.48 | 8.01 | -3.09, 7.42 | -0.24 [-1.50, 1.01] | Number of prior injuries, Baseline VO_2_max, total met |
| **Cardiorespiratory Fitness** | 185.52 | -0.05 [-1.20, 1.11] | 179.8 | 5.71 | -6.91, 5.45 | -0.77 [-2.06, 0.52] | Baseline symptoms, time in heart rate |
| **Concussion Symptoms** | 84.15 | -0.48 [-1.67, 0.72] | 81.63 | 2.51 | -0.75, 0.67 | -0.64 [-1.85, 0.56] | Total met |

Supplementary Material 13: Group x Time results

Group x Time Interaction for Primary outcomes

A Group×Time interaction for HVLT delayed recall (β=1.50,SE=0.74,g=1.15,95%CI[0.03,2.26]), indicating that the two groups showed different patterns of change in delayed word recall over the course of the intervention. The interaction effect represents a large effect size (g=1.15), suggesting that the trajectory of improvement in delayed word recall differed meaningfully between the aerobic exercise and balance training groups (Figure 4e). The aerobic group had a negative slope [β=-0.5,95%CI(-1.53, 0.53)], and the balance group had a positive slope [β=1.0,95%CI(-0.03,2.03)], suggesting the balance group improved over the 12 weeks. A Group×Time interaction for TMT B-A (β=0.62,SE=1.50,g=1.38,95%CI[0.27,2.49]), indicating that the two groups showed different patterns of change in TMT B-A over the course of the intervention (Figure 4f). The interaction effect represents a large effect size (g=1.38), suggesting that the trajectory of improvement in delayed verbal recall differed meaningfully between the aerobic exercise and balance training groups. The aerobic group had a negative slope [β=-10.5,95%CI(-17.35,-3.65)], and the balance group had a positive slope [β=1.50,95%CI(-5.35,8.35)], suggesting the aerobic group got faster over the 12 weeks.

The analysis of HVLT delayed word recall revealed a Group × Time interaction, indicating different cognitive recovery trajectories between groups. However, interpretation requires careful consideration of baseline differences and regression to the mean. The aerobic group began with higher scores (M = 10.75) compared to the balance group (M = 8.92), creating conditions where measurements naturally trend toward the population average upon repeated assessment. By post-intervention, the groups converged to nearly equivalent performance (aerobic: M = 10.25, balance: M = 9.92), with no between-group differences, though the ANCOVA yielded a large effect size with wide confidence intervals reflecting considerable uncertainty. While regression to the mean provides a parsimonious explanation for these trajectories, alternative mechanisms warrant consideration. The balance group's improvement may reflect genuine therapeutic benefit through engagement of cerebellar and vestibular systems involved in memory encoding^1^, while both interventions provided structured activity and cognitive engagement that may support recovery. Additionally, the integration of multisensory information and dual tasking during balance training might engage distributed neural networks that overlap with memory circuitry^2^, explaining why balance interventions showed unexpected benefits for recall despite their lower cardiovascular demands. Supporting this, a prior study in healthy older adults demonstrated that balance training benefited memory but not executive functioning^28^. Speculatively, different types of physical exercise are important for different cognitive domains, thus encouraging precision medicine approach to identify which therapeutic intervention is needed based on individual deficits. Nevertheless, these findings underscore the importance of balanced baseline cognitive performance in future trials and suggest that stratified randomization procedures, additional measurement timepoints, and adequately powered samples would enable clearer interpretation of treatment effects on memory outcomes following mTBI.

Group x Time Interaction

| **Measure** | **Base AIC** | **Base Hedges' g (95% CI)** | **Final AIC** | **AIC Change** | **β, SE** | **Final Hedges' g (95% CI)** | **Optional Covariates** |
| --- | --- | --- | --- | --- | --- | --- | --- |
| **Hopkin Verbal Learning Test Total Recall** | 284.82 | 0.86 [-0.25, 1.97] | 283.310866 | 1.51 | 3.17, 2.09 | 0.86 [-0.25, 1.97] | time in heart rate |
| **Hopkin Verbal Learning Test Delayed Recall** | 213.61 | 1.15 [0.03, 2.26] | 213.607686 | 0 | 1.50, 0.74 | 1.15 [0.03, 2.26] | None |
| **Hopkin Verbal Learning Test Retention** | 14.63 | 0.38 [-0.74, 1.49] | 14.6307212 | 0 | 0.05, 0.07 | 0.38 [-0.74, 1.49] | None |
| **Hopkin Verbal Learning Test Recognition Discrimination Index** | 163.48 | 0.75 [-0.37, 1.86] | 163.479898 | 0 | 0.58, 0.44 | 0.75 [-0.37, 1.86] | None |
| **Verbal Fluency Letter** | 348.54 | -0.78 [-1.89, 0.34] | 344.16463 | 4.38 | -4.75, 3.48 | -0.78 [-1.89, 0.34] | Number of prior injuries, time in heart rate |
| **Verbal Fluency Animals** | 278.52 | -0.07 [-1.19, 1.04] | 276.947645 | 1.58 | -0.25, 1.91 | -0.07 [-1.19, 1.04] | time in heart rate |
| **Trail Making A** | 303.15 | -0.68 [-1.79, 0.43] | 300.821612 | 2.32 | -2.5, 2.09 | -0.68 [-1.79, 0.43] | time in heart rate |
| **Trail Making B** | 393.63 | 1.07 [-0.04, 2.19] | 385.98524 | 7.64 | 9.5, 5.03 | 1.07 [-0.04, 2.19] | Number of prior injuries, Adherence Rate, time in heart rate |
| **Trail Making B-A** | 388.69 | 1.38 [0.27, 2.49] | 386.31893 | 2.37 | 12.00, 4.94 | 1.38 [0.27, 2.49] | Number of prior injuries, Adherence Rate |
| **Pittsburg Sleep Quality Index Global** | 224.22 | 0.25 [-0.95, 1.45] | 223.743913 | 0.48 | 0.62, 1.50 | 0.26 [-0.95, 1.46] | time in heart rate |
| **PROMIS Global Physical Health T-Score** | 338.16 | -0.51 [-1.62, 0.61] | 328.9 | 9.19 | -3.167, 3.559 | -0.51 [-1.62, 0.61] | Number of prior injuries, time in heart rate |
| **PROMIS Global Mental Health T-Score** | 348.64 | 0.20 [-0.92, 1.31] | 339.506692 | 9.13 | 1.46, 4.17 | 0.20 [-0.92, 1.31] | Number of prior injuries, time in heart rate |
| **PROMIS Cognitive T-Score** | 355.0346 | 0.031 [-1.082, 1.145 ] | 349.1382 | 5.9 | 0.225, 4.085 | 0.031 [-1.082, 1.145 ] | Number of prior injuries, time in heart rate |
| **Balance Error Scoring System** | 312.78 | 1.00 [-0.12, 2.11] | 309.686929 | 3.09 | 4.5. 2.56 | 1.00 [-0.12, 2.11] | time in heart rate |
| **Resting Heart Rate** | 380.56 | 0.01 [-1.11, 1.12] | 366.903281 | 13.66 | 0.08, 5.87 | 0.01 [-1.11, 1.12] | Number of prior injuries Baseline VO_2_max,, time in heart rate |
| **Cardiorespiratory Fitness** | 346.71 | 0.02 [-1.10, 1.13] | 342.813797 | 3.89 | 0.11, 3.91 | 0.02 [-1.10, 1.13] | time in heart rate |
| **Concussion Symptoms** | 361.15 | 0.67 [-0.44, 1.78] | 354.320814 | 6.83 | 6.75, 5.73 | 0.67 [-0.44, 1.78] | Number of prior injuries, time in heart rate |

Supplementary Material 14: Intervention feasibility

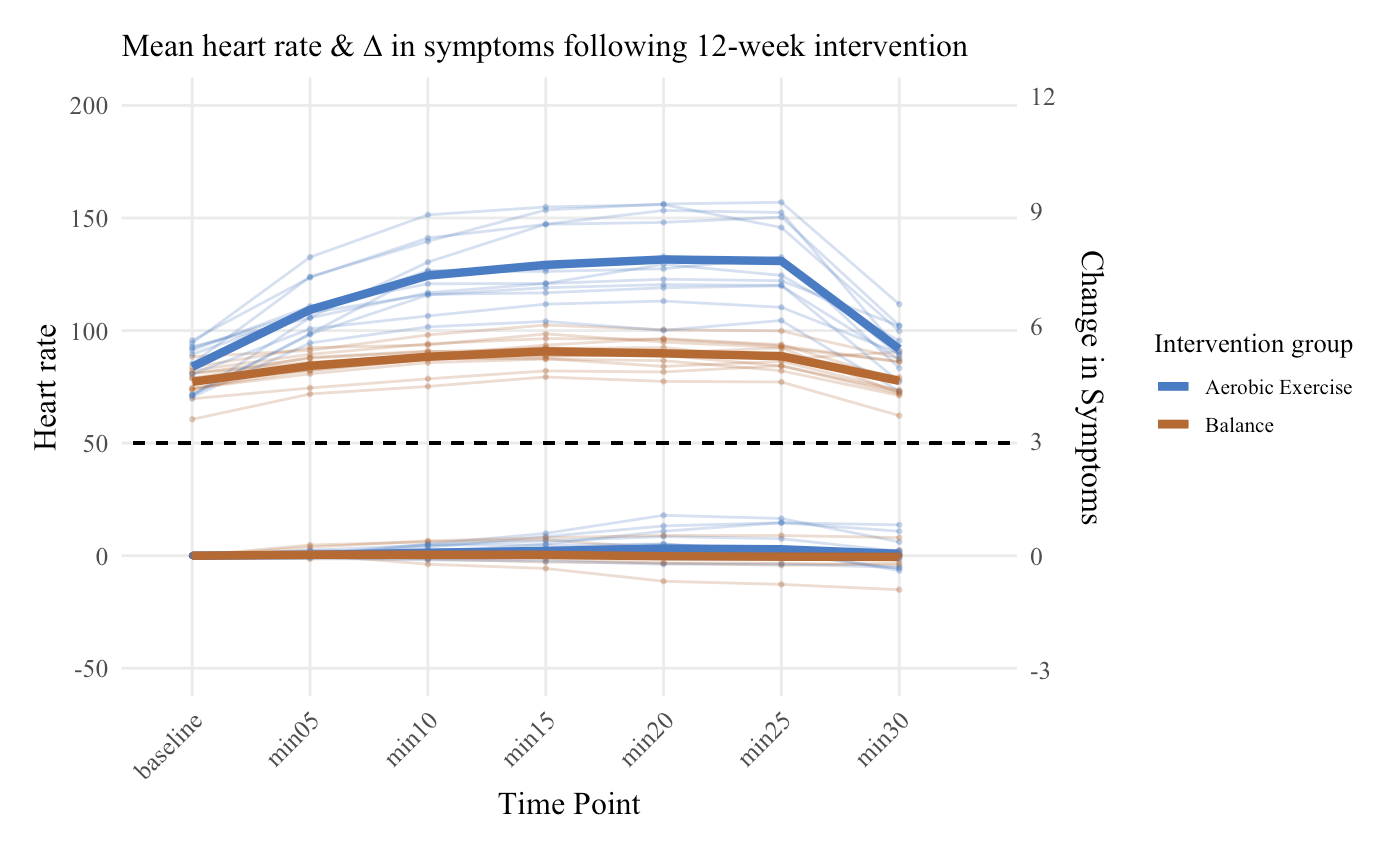

Figure 14: Mean Heart Rate and Change in symptoms over 12-week intervention

1.	Besnard S, Lopez C, Brandt T, Denise P, Smith PF. Editorial: The Vestibular System in Cognitive and Memory Processes in Mammalians. *Front Integr Neurosci*. 2015;9. doi:10.3389/fnint.2015.00055

2.	Surgent OJ, Dadalko OI, Pickett KA, Travers BG. Balance and the brain: A review of structural brain correlates of postural balance and balance training in humans. *Gait Posture*. 2019;71:245-252. doi:10.1016/j.gaitpost.2019.05.011

3.	Rogge AK, Röder B, Zech A, et al. Balance training improves memory and spatial cognition in healthy adults. *Sci Rep*. 2017;7(1):5661. doi:10.1038/s41598-017-06071-9
